# Your body, my experience: A systematic review of embodiment illusions as a function of and method to improve body image disturbance

**DOI:** 10.1101/2023.10.21.23297282

**Authors:** Jade Portingale, Isabel Krug, Hermione Liu, Litza Kiropoulos, David Butler

## Abstract

Embodiment illusions—which manipulate multisensory integration processes underlying self-perception—are increasingly used to understand and improve body image disturbance (BID): a core symptom of eating disorders (EDs) and body dysmorphic disorder (BDD). The current systematic review is the first to evaluate clinical and community-based evidence on whether (i) variations involving BID impact embodiment illusion susceptibility and (ii) embodiment illusions can improve BID. Thirty-two studies met inclusion criteria. Greater embodiment was generally observed among individuals with high (relative to low/no) BID (9 out of 14 studies; 64.29%). Embodiment produced an improvement in BID in most studies (20 out of 24; 83.33%). Effect sizes were generally medium to large across both findings. Although several issues exist within the literature (e.g., substantial methodological heterogeneity, non-validated measures), findings reiterate that disturbances in multisensory integration appear to underpin BID, although embodiment illusions offer opportunities to develop new therapeutic interventions for BID. Multiple factors are of particular interest for ensuring that embodiment illusion paradigms are best designed for future research and application (e.g., virtual reality versus real-world methods).

## Introduction

Body image disturbance (BID) is a multidimensional concept characterised by numerous components (American Psychiatric Association [APA], 2013). The most widely accepted components are cognitive-affective (i.e., dysfunctional negative thoughts, beliefs, and feelings towards body weight or shape; e.g., body dissatisfaction) and perceptual (i.e., distorted experience of body weight or shape; e.g., body size overestimation or underestimation; Hosseini & Padhy, 2019). BID is a central factor in the onset, maintenance, and relapse of eating disorders (EDs; Glashouwer et al., 2019) and body dysmorphic disorder (BDD; Mitchison & Mond, 2015).

Historically, research has predominately focused on cognitive-affective BID within EDs (Urgesi, 2015) and BDD (Kaplan et al., 2014), with current best practice treatments also commonly targeting this dimension (for review see Aleva et al., 2015). Perceptual bodily disturbance remains comparably less understood despite contributing to poorer clinical outcomes than the cognitive-affective dimension (Beohm et al., 2016; Eshkevari et al., 2012). Overall, this warrants continuing research to increase understanding of the mechanisms underpinning BID and its effective treatment. The current review will investigate whether and how a self-perceptual phenomenon known as illusory embodiment can be used to understand the basis of and potentially improve BID.

### Multisensory integration and bodily-self-perception

Bodily self-perception results from combining real-time sensory inputs from two or more modalities, involving exteroception (i.e., visual and tactile inputs), interoception (i.e., internal body input), and proprioception (i.e., the sense of body position/movement in space; Brizzi et al., 2023). Multisensory integration enables us to continually update how we perceive and recognise ourselves (Longo et al., 2008), as evidenced via multisensory bodily (embodiment) illusions (Kilteni et al., 2015).

For instance, the classic rubber hand illusion (RHI; Figure 1) involves a three-way interaction between vision (i.e., seeing a rubber hand stroked), touch (i.e., feeling one’s [unseen] hand receive stroking), and proprioception (i.e., sensing the spatial location of one’s hand; Botvinick & Cohen, 1998). Synchronous (but typically not asynchronous) interpersonal multisensory stimulation involving both hands generally produces the temporary illusorily experience of ‘embodying’ the artificial hand (Longo et al., 2008).

**Figure 1.**
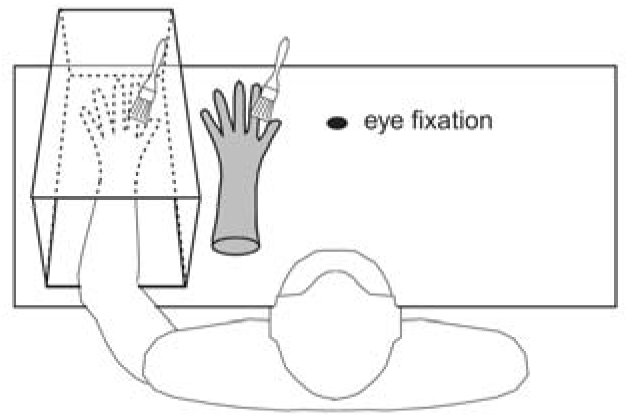
Illustration of the ‘Classic’ Rubber Hand Illusion. *Note.* A participant’s hand is obscured from view while a rubber hand is placed in view. Both hands are simultaneously stroked. Image used with permission from Mussap & Salton (2006).

‘Full-body’ and ‘enfacement’ illusions—which employ the same multisensory principles as the RHI to one’s whole body and face, respectively—have also been developed. Full-body illusions (see Figure 2) are typically generated using immersive virtual reality (VR). Whilst wearing a head-mounted display, participants observe a virtual body (avatar) that visually substitutes their own (unseen) body from a first-person perspective (Normand et al., 2011; Piryankova et al., 2014). Visuo-tactile (i.e., touches or vibrations delivered to the participant’s and avatar’s corresponding body parts) or visuo-motor (i.e., the participant’s and avatar’s movements correspond) stimulation can induce embodiment.

**Figure 2.**
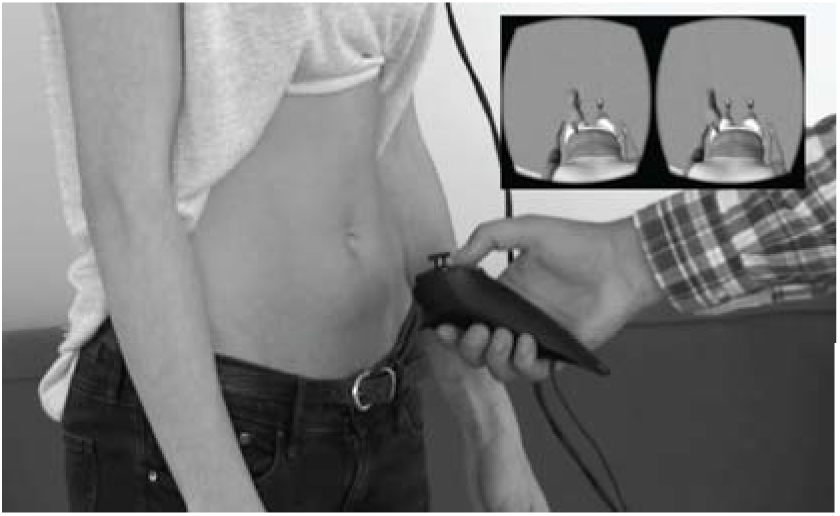
Example of the Procedure Used to Induce the Full-Body Illusion in a Virtual Reality Setting. *Note.* Participant observes an avatar’s abdomen being stroked synchronously with their own abdomen from a first-person perspective via a head-mounted display (top right image). Tactile stimulation is administered by the experimenter (main image). Figure used with permission from Keizer et al., 2016.

Enfacement illusions typically involve watching another person’s/model’s face (e.g., via a computer screen or sitting opposite) whilst experiencing visuo-motor (i.e., participants mimic the actor’s facial expressions) or visuo-tactile (i.e., participant’s and actor’s faces are stroked with a brush or cotton tip) stimulation (e.g., Ma et al., 2016).

Embodiment illusions are typically measured subjectively via self-report questionnaires assessing ownership, agency, and self-location over the other body part, and objectively via perceptual or behavioural tasks (e.g., body size estimation). Please see Appendix A for a full summary of measures.

Embodiment illusions are generally interpreted within a predictive coding framework of brain functioning (for review see Tsakiris, 2017). This theory proposes that *current* self-perception relies on the brain consistently interpreting lower-level (bottom-up) sensory information under a higher-level (top-down) predictive model of what is most likely to be ‘self’ based on *past* perceptual experiences (for review see Apps & Tsakiris, 2013). Incongruence between the predicted and actual sensory input generates prediction errors which are reconciled by re-aligning the incoming signals with the predictive model. For example, in the RHI, the brain’s initial predictive model of the rubber hand as ‘not-self’ based on internal proprioceptive and interoceptive information (“*my hand is placed under the box, not where the rubber hand is*”), becomes incongruent with incoming external visual and tactile input (“*I feel my hand receive stroking as I see the rubber hand receive the same”*). This predictive error is reconciled by updating the model to now consider the rubber hand as one’s own (Tsakiris, 2017). Whether and how these theoretical principles shed light on distorted self-perception among individuals with BID will now be considered.

### Embodiment illusions in individuals with BID

Prior research has revealed higher-order perceptual disturbances amongst individuals with clinically significant (i.e., EDs, BDD) and sub-threshold BID across visual, tactile (Engel & Keizer, 2017), proprioceptive (Eshkevari et al., 2012), and interoceptive (Badoud & Tsakiris, 2017) sensory domains. Researchers have increasingly begun to use embodiment illusions to investigate whether and how disturbances in multisensory integration influence body perception in clinical populations (for reviews see Moseley et al., 2012; Matamala-Gomez et al., 2021). To our knowledge, only one non-systematic review by Crespi and Dinsdale (2019) has examined susceptibility to embodiment illusions across a range of mental disorders and conditions: specifically, those believed to experience either heightened (e.g., EDs, BDD, psychotic disorders) or reduced susceptibility (e.g., autism spectrum disorder) to embodiment. Interestingly, five (71%) out of seven ED/BDD-based studies observed greater susceptibility among those with high (relative to low/no) symptoms. However, their review was restricted to the RHI, thus, failing to capture studies examining other embodiment paradigms. No researchers have systematically reviewed the literature regarding susceptibility to embodiment illusions as a function of BID.

### Using embodiment illusions to improve BID

Although evidence-based treatments for EDs and BDD have significantly improved in quality and quantity over recent years, they often achieve modest results (for review see Ziser et al., 2018). For instance, the widely accepted cognitive behavioural therapy (CBT; Hay, 2020) typically focuses on weight restoration or minimising cognitive-affective or behavioural symptoms (e.g., fear of weight gain, loss of control eating; Kass et al., 2013). Generally, only 30-50% of ED patients report cessation of symptoms following CBT, with the remaining exhibiting partial remission, no or minimal improvement, or premature withdrawal from treatment (Atwood & Friedman, 2020). Similarly, typically only 40-50% of BDD patients respond to CBT (Öst et al., 2019), with many requiring long-term treatment (Flygare et al., 2020).

Intervention strategies targeting distorted self-perception are less common than those targeting cognitive-affective or behavioural symptoms and the efficacy of current perceptual approaches (e.g., mirror exposure therapy) remains unclear (for review see Ziser et al., 2018). Effective strategies targeting distorted body perception are required given that these distortions are a risk factor for cognitive-affective disturbance (Keizer et al., 2014) and are highly associated with ED (Engel & Keizer, 2017) and BDD (Khemlani-Patel et al., 2011) maintenance and relapse.

Embodiment illusions target the processes underlying self-perception and demonstrate how it is a dynamic and malleable construct. Their potential benefit is that the amalgamation of the embodiment model into one’s self-representation can be used to temporarily update, and thus improve BID (Keizer et al., 2016; Preston & Ehrsson, 2014). That is, one may be able to see their features more positively (e.g., torso width or some other perceived defect) due to the assimilation of the more socio-culturally desirable features of the embodiment model. Given the general tendency for individuals with BID to overestimate their body size (Gardner & Brown, 2014), reductions in body size estimation (and misestimation) could be considered an improvement.

Prior non-systematic reviews have provided a clinical rationale for the use of embodiment illusions in mental health (Moseley et al., 2012), and importantly, researchers have increasingly investigated the effects of embodiment illusions on improving BID. Earlier non-systematic reviews (Matamala-Gomez et al., 2021, Riva et al., 2021) have broadly explored and supported the potential efficacy of embodiment illusions, specifically full-body illusions, for improving BID. To date, only one systematic review and meta-analysis by Turbyne et al. (2021) has evaluated the efficacy of VR-based full-body illusions in improving BID. Of the 13 reviewed studies, nine (69%) found improvements in BID post-embodiment. However, their review was restricted to full-body illusions and ‘body image’ constructs, thus failing to capture studies assessing other embodiment paradigms and ED-specific outcomes (e.g., fear of gaining weight). Moreover, the authors conceptualised *improvement* according to body dissatisfaction, neglecting perceptual BID. These limitations must be addressed before concluding the efficacy of using these illusions in BID populations.

### Additional influencing factors

Several possible factors warrant consideration when systematically reviewing the BID literature assessing susceptibility to embodiment illusions and post-embodiment improvements. First, although BDD and EDs (including their sub-types and symptoms) share core features, from a current clinical perspective, they exhibit distinct manifestations (APA, 2013) and should be examined independently.

Second, the effects of real-world versus VR-based embodiment illusion paradigms among BID populations remain unclear and may differ; for instance, depending on brain responsiveness to different stimuli or the customisability of stimuli enabled by the paradigm.

Third, scant research has directly compared the effect of the body part (or the number of parts) stimulated in BID populations. Differences may emerge based on its cognitive-affective salience (Frederick et al., 2016) or the density of sensory receptors reflecting that region within the brain (Corniani & Saal, 2020).

Fourth, whilst embodiment has been argued to emerge regardless of the model’s visual size (Normand et al., 2011; Preston & Ehrsson, 2014), the size of the embodied body part relative to one’s body size (i.e., smaller, larger, or same-sized) may influence improvements in BID post-embodiment. Turbyne et al.’s review (2021) reported that embodying a smaller (relative to larger) body leads to greater improvements in BID: however, conclusions remain tentative as their findings were based on VR-based full-body illusions alone and the effects of embodying same-sized bodies are unclear.

Finally, embodiment illusions are typically sensitive to the synchronous temporal aspects of visual and tactile stimulation, with asynchronous stimulation typically used as a control condition (Longo et al., 2008). However, it is unclear whether the effects of temporal synchrony during interpersonal multisensory stimulation in BID populations may manifest differently. Turbyne et al.’s (2021) review found that individuals with BID still experienced embodiment and changes in BID post-asynchronous stimulation, though to a lesser extent than post-synchronous stimulation.

### The current systematic review

Despite the growing number of studies, there has been no attempt to systematically review the embodiment illusion literature concerning both susceptibility and effectiveness as a function of BID. The present systematic review has two primary aims. First, to examine whether susceptibility to embodiment illusions differs as a function of varying levels of BID. This may better elucidate whether and/or how individuals with high BID exhibit specific deficits/distortions in self-perception. Second, to assess whether embodiment illusions can improve BID. This may assist in developing alternative prevention, early intervention, and treatment approaches targeting EDs and BDD. We will also explore whether additional population-related factors (ED or BDD diagnosis) and embodiment-related phenomena (e.g., body part targeted) impact susceptibility and improvements to enhance insight into how these illusions can be most effectively conducted.

We hypothesise that (1) individuals with high (relative to low/no) BID will be more susceptible to embodiment illusions, and (2) individuals (irrespective of pre-existing BID) will show improvements in BID after experiencing embodiment illusions.

## Method

### Selection of studies

The primary aims and methods of the current systematic review were pre-registered in the PROSPERO database (ID CRD42021277687). The methodology for the present review followed the Preferred Reporting for Systematic Reviews and Meta-Analyses (PRISMA) guidelines (Moher et al., 2009). Relevant studies were identified by searching the international online databases PsycINFO, MEDLINE, EMBASE, Web of Science, and CINAHL. ProQuest Dissertations and Theses Global were also searched to identify relevant unpublished (grey) literature. Citation searching of included articles was also conducted. An initial search was conducted on August 25, 2021, and then updated on May 25, 2022, and December 23, 2022, following identical search and coding procedures.

For each database, keywords (and related terms) from the embodiment illusion literature were entered into the field of keyword, title, and/or abstract. They were combined with additional terms (synonyms) related to the key concepts of (1) body image, (2) eating symptomatology, and (3) dysmorphic symptomatology, using the conjunction “AND” and separate searches for each combination. The search syntax was created by the first author J. P. and checked by a librarian. Please see Appendix B for the full search syntax.

The inclusion criteria were studies that examined and measured (a) at least one multisensory bodily illusory paradigm (e.g., the RHI) concerning (b) at least one BID-related variable. This included (i) clinical EDs or BDDs (e.g., anorexia nervosa, bulimia nervosa, muscle dysmorphia) or (ii) BID-related symptoms or obesity (as a means of representing BID; for review see Weinberger et al., 2017) within a community-based sample. *Improvement* in BID was considered perceptual (conceptualised as reduced body size estimation or misestimation relative to baseline) or cognitive-affective (conceptualised as an improved rating on a BID-related scale relative to baseline). Studies that assessed *changes* in BID were included.

Exclusion criteria included: (a) studies that examined embodiment illusions and BID exclusively among clinical populations or conditions that are not of central interest to this review—e.g., schizophrenia, depression, physical condition/injury, or neurological conditions (e.g., epilepsy); (b) studies that exclusively employed neurobiological outcome measures (e.g., functional magnetic resonance imaging [fMRI]); (c) failure to draw conclusions based upon inferential statistics involving quantitative data (i.e., significance testing based upon *p* < .05 or adjusted levels if multiple comparisons); (d) non-empirical studies (e.g., reviews, case studies); (e) qualitative studies; (f) and if the full-text was unavailable.

Database searches identified 5,115 potential studies. The Covidence platform (Veritas Health Innovation, 2022) aided in screening and selection. Titles and abstracts were screened against the inclusion and exclusion criteria by J. P. and independently by a second reviewer (K. P.). Full texts of all remaining articles were examined independently by J. P. and K. P. to determine their eligibility, with no discrepancies between reviewers. After removing duplicates, 1,555 studies were screened via titles and abstracts, and then 99 full-text records were assessed for eligibility: 69 were excluded as they did not meet the criteria, leaving a final total of 32 studies. Figure 4 summarises the selection process and reasons for exclusion.

**Figure 4.**
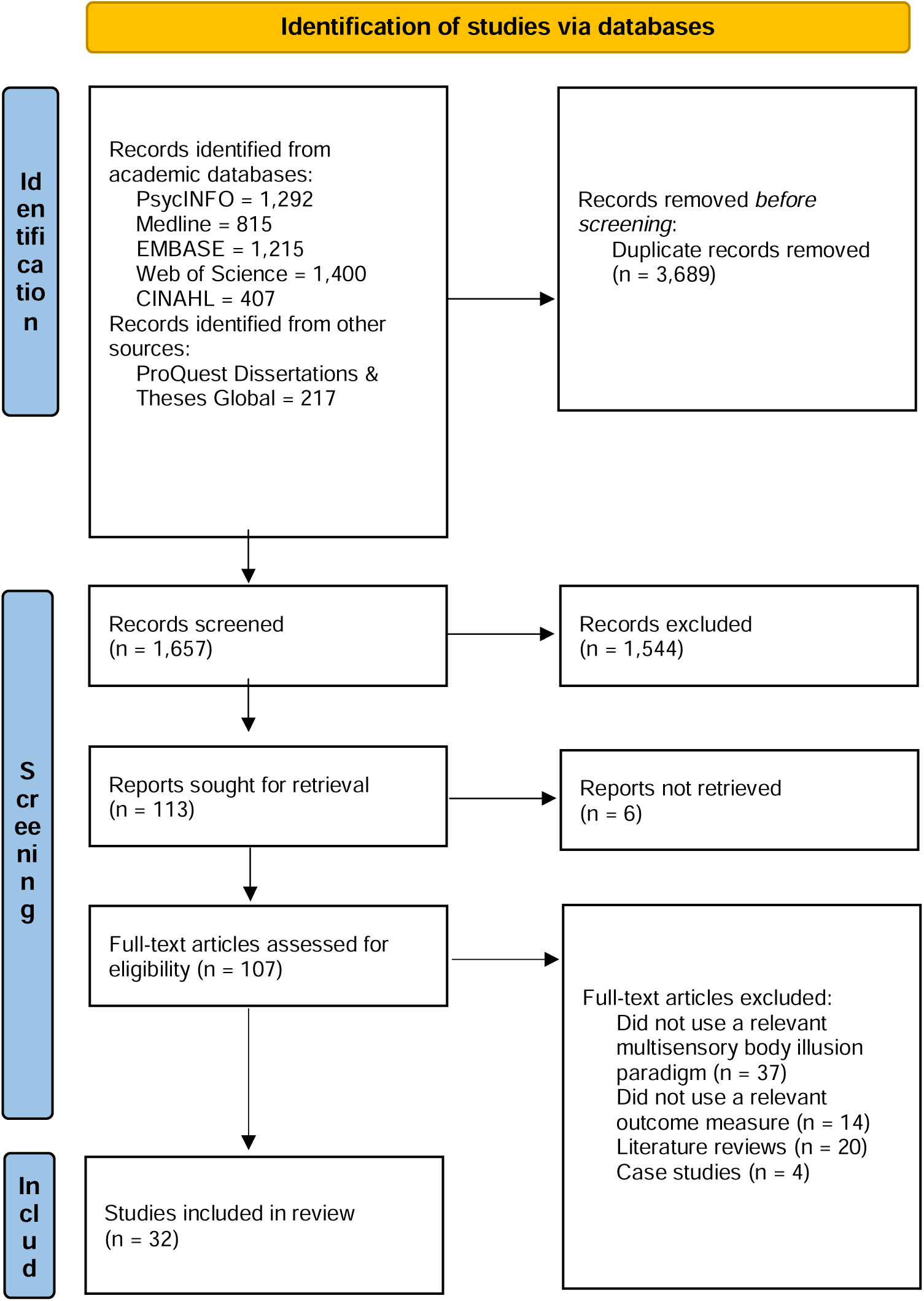
PRISMA Flow Chart of Systematic Review Selection Process. *Note.* Adapted PRISMA 2020 flow diagram for systematic reviews which include searches of databases and registers only.

### Data extraction

Extracted data for the 32 studies included: (a) bibliographical data (author names, publication year); (b) aim(s) (as relevant to the current systematic review); (c) study details (country, setting; e.g., clinical, community); (d) sample and descriptive statistics (sample size, age, body mass index [BMI], gender, clinical diagnosis); (e) stimuli and measures (as relevant to the current systematic review); (f) procedure; (g) key findings (addressing the aims of the current systematic review) based upon statistical significance and effect size; and (h) strengths and limitations. Relevant data were extracted by J. P. using Covidence and cross-checked by H. L. Please see Appendix C for the summary of results table.

Included studies had significant statistical, methodological, and measurement heterogeneity. Rather than attempting a meta-analysis, we undertook a systematic narrative synthesis of results using (i) a vote count approach (i.e., X out of Y studies) based upon inferential statistics and (ii) effect size (Higgins et al., 2019; Moher et al., 2009). Cohen’s *d* was chosen as the measure of effect size, with effects of 0.2, 0.5, and 0.8 deemed small, medium, and large, respectively (Cohen, 1988). If Cohen’s *d* was not reported and relevant data was available (e.g., means and standard deviations, t-tests, or analysis of variance), then it was calculated using the online calculator Psychometrica (Lenhard, 2016).

### Quality appraisal and risk of bias assessment

Methodological quality and risk of bias were assessed using Rozenblat et al.’s (2017) revised version of Downs and Black’s (1998) original criteria, adapted for the BID-related literature. Thirteen criteria assessed whether studies (a) clearly described the aims, objectives, and hypotheses; (b) clearly outlined the main interventions and/or measures in the introduction or method; (c) specified participant characteristics; (d) clearly described interventions of interest; (e) clearly described principal confounders; (f) stated characteristics of participants lost at follow-up (if relevant); (g) clearly described the main findings; (h) included exact probability values (or confidence intervals included); (i) used a representative sample of the population examined (i.e., including clinical but not convenience samples); (j) noted any “data-dredging” explicitly; (k) used appropriate statistical tests; (l) employed valid and reliable main outcome measures; (m) conducted adequate adjustment for confounding variables; and (n) had sufficient power. There was a maximum score of 15 points^1^ [^1^One criterion (e) carried a possible two points. The power question was simplified to a possible one-point based on prior recommendations.]. Using a modified grading system (O’Connor et al., 2015), scores were converted to percentages, then studies were assigned a grade of “excellent” (>86%), “good” (85%-71%), “fair” (70%-50%) or “poor” (<50%). Studies were evaluated independently by two coders (J. P., H. L.) and then cross-checked, with no concerns raised.

## Results

### Study characteristics

The main characteristics and results of the 32 included studies are summarised in Appendix C. Studies were published between the years 2006 and 2022 in the English language. Thirty studies were published in peer-reviewed journals and two were from the grey literature (Lavenne-Collot et al., 2022; Wolf et al., 2021). Sample sizes ranged from six to 167, with a median of 36. The total participants numbered 1,640, of which 1,339 (81.65%) were female (382 from ED samples, 13 from a BDD sample, and 944 from community-based samples or healthy control groups), 260 (15.85%) were male (five from an ED sample, four from a BDD sample, and 251 from community-based samples or healthy control groups), and 41 (2.50%) were unidentified regarding gender due to an absence of information. Nineteen studies examined females exclusively, ten examined a mixed-gender sample, two examined males exclusively, and one was gender-unidentified. The mean age of participants ranged from 17 to 37 years, though the dominant age group was early 20s. The mean BMI (kg/m^2^) ranged from 15.50 to 19.80 for ED samples (reflecting the various ED subtypes) and from 18.98 to 45.33 for community-based samples (the upper range reflected two studies purposely assessing obese samples). Mean BMI was not reported for the BDD sample.

Most studies were European, being conducted in Italy (*n* = 8), Spain (*n* = 7), Germany, the United Kingdom (*n*s= 3), France, Sweden, and the Netherlands (*n*s = 2). Non-European countries included Australia (*n* = 4) and Taiwan (*n* = 1). Participants with EDs or BDD were generally drawn from voluntary ED databases or body image/ED clinics in hospital settings. Most community-based samples and healthy controls were recruited through convenience sampling (e.g., university laboratories) or via community advertisement.

Regarding samples, 13 studies examined clinical conditions: specifically, anorexia nervosa (*n* = 9), a mixed ED group (*n* = 3; i.e., anorexia nervosa, bulimia nervosa, binge eating disorder, and other specified feeding and eating disorder), and BDD (*n* = 1). Of these studies, 31% verified diagnoses via structured/semi-structured interviews following DSM-IV (Eshkevari et al., 2012; Keizer et al., 2016), DSM-IV-TR (Kaplan et al., 2014) or DSM-V criteria (Eshkevari et al., 2014); the remaining relied on current DSM-V diagnoses made externally (e.g., within hospitals) without interview-based verification. Nine of these 13 studies utilised healthy control samples, however, only 38% of these samples were screened for BID using validated measures (e.g., Eshkevari et al., 2012; 2014). The remaining 19 studies examined a community-based population: two compared individuals with obesity (i.e., likely sub-threshold BID) to healthy-weight controls, though neither screened for BID in the latter (Scarpina et al., 2019; Tagini et al., 2020). Research designs were predominately quasi-experimental, with three studies having a longitudinal design.

### Synthesis of main findings

#### Aim 1. Susceptibility to embodiment illusions as a function of BID

Fourteen studies examined differences in susceptibility to embodiment illusions as a function of BID. Nine (64.29%) reported significantly greater susceptibility amongst those with high (relative to low/no) BID on at least one outcome measure (generally involving medium or large-sized effects; Eshkevari et al., 2012; 2014; Kaplan et al., 2014; Keizer et al., 2014; 2016; Lavenne-Collot et al., 2022; Metral et al., 2017; Mussap & Salton, 2006; Zopf et al., 2016). Three (21.43%) studies reported reduced susceptibility among those with high (relative to low/no) BID on at least one measure, involving medium and large effects (Carey & Preston, 2019; Porras-Garcia et al., 2020; Tagini et al., 2020). The remaining two (14.49%) observed no significant effect of BID on embodiment: although effect sizes ranged from negligible to medium, though, the direction of effects was not discernible (Provenzano et al., 2019; Scarpina et al., 2019).

**1a. Clinical conditions and symptoms.** Of the 14 studies, six compared individuals with anorexia nervosa to healthy controls: of these, four (66.67%) reported greater susceptibility in the anorexia sample (generally involving medium or large effects; Keizer et al., 2014; 2016; Lavenne-Collot et al., 2022; Zopf et al., 2016); one reported greater susceptibility among the healthy control group (with a medium effect; Porras-Garcia et al., 2020); and the remaining study found no significant difference (with a not discernible small-sized effect; Provenzano et al., 2019). Three studies compared a mixed ED group to healthy controls: two (66.67%) reported greater susceptibility among the ED group, with small to large-sized effects (Eshkevari et al., 2012; 2014). The remaining study reported no significant between-group difference (although negligible to small-sized effects favouring greater embodiment among the ED group were evident; Carey & Preston, 2019). One study compared BDD individuals to healthy controls and reported no significant difference (although a medium-sized effect favouring greater embodiment among those with BDD was evident). However, significant medium and large-sized positive associations were found between BID-related symptoms (e.g., bulimia, body dissatisfaction) and susceptibility across the entire sample (Kaplan et al., 2014).

Four studies assessed a community-based population. Two used a dimensional approach and reported significant medium or large-sized positive associations between embodiment susceptibility and BID symptoms (e.g., binge eating/purging, frequent chemical supplement usage; Metral et al., 2017; Mussap & Salton, 2006). The remaining two studies compared those with obesity to healthy controls: one reported greater susceptibility among healthy controls (large-sized effect; Tagini et al., 2020) and the other found no significant between-group difference (with a not discernible negligible effect; Scarpina et al., 2019).

**1b. Embodiment illusion paradigm.** Of the 14 studies, nine employed a real-world paradigm (RHI [*n* = 8]; double-mirror paradigm [*n* = 1]), of which, eight (88.89%) reported greater susceptibility for those with high (relative to low/no) BID (generally involving medium to large-sized effects; Eshkevari et al., 2012; 2014; Kaplan et al., 2014; Keizer et al., 2014; Lavenne-Collot et al., 2022; Metral et al., 2017; Mussap & Salton, 2006; Zopf et al., 2016). The remaining study reported reduced susceptibility among those with high BID (medium-sized effect; Carey & Preston, 2019). Five studies utilised a VR-based paradigm (full-body illusion [*n* = 4]; virtual-hand illusion [*n* = 1]): one (20.0%) observed greater susceptibility for those with high BID (small to large-sized effects; Keizer et al., 2016), two observed reduced susceptibility for those with high BID (medium to large-sized effects; Porras-Garcia et al., 2020; Tagini et al., 2020), and two observed no significant differences (with negligible to medium-sized effects in a non-discernible direction^2^ [^2^The term ‘non-discernible’ refers to the *direction* of the effect for non-significant findings.]; Provenzano et al., 2019; Scarpina et al., 2019).

**1c. Body part(s) embodied.** Of the 14 studies, nine stimulated the hand alone, of which, seven (77.78%) reported greater susceptibility for those with high (relative to low/no) BID (generally medium to large-sized effects; Eshkevari et al., 2012; 2014; Kaplan et al., 2014; Keizer et al., 2014; Metral et al., 2017; Mussap & Salton, 2006; Zopf et al., 2016). Of the remaining hand-based studies, one observed greater susceptibility among those with no (relative to likely high) BID (large-sized effect; Tagini et al., 2020), whilst another reported no significant effect of BID on embodiment (although there were negligible to medium effects in the expected direction; Carey & Preston, 2019). One study utilised the face and reported greater susceptibility among those with high (relative to low/no) BID (large-sized effects; Lavenne-Collot et al., 2022). Three studies stimulated the abdomen (stomach/torso/belly) alone: one (33.33%) observed greater susceptibility among those with high (relative to low/no) BID (with small to large-sized effects; Keizer et al., 2016), and two reported no significant effect of BID on embodiment (with non-discernible negligible to medium-sized effects; Provenzano et al., 2019; Scarpina et al., 2019). The remaining study stimulated the abdomen, and upper and lower limbs and reported reduced susceptibility for those with high BID (with a medium-sized effect; Porras-Garcia et al., 2020).

**1d. Synchrony of stimulation**. Of the nine studies that reported greater susceptibility among those with high (relative to low/no) BID, six compared synchronous versus asynchronous stimulation. Five (83.33%) observed the effect irrespective of synchrony (generally medium to large-sized effects), though synchronous stimulation produced stronger effects when compared (large-sized effects; Eshkevari et al., 2012; 2014; Kaplan et al., 2014; Keizer et al., 2016; Zopf et al., 2016). Another study observed a significant medium-sized effect only in synchronous conditions, though noting a non-significant small-sized effect for asynchronous conditions (Keizer et al., 2014). The remaining three studies failed to include an asynchronous condition (Mussap & Salton, 2006), compare the two during analyses (Metral et al., 2017), or assess synchrony (i.e., mere observation of another body part with no stimulation; Lavenne-Collot et al., 2022).

#### Aim 2. Embodiment illusions improving BID

Twenty-four studies examined the effect of embodiment illusions on BID using perceptual and/or cognitive-affective measures. Twenty (83.33%) observed small to large-sized (predominately medium to large-sized) improvements post-embodiment (Carey & Preston, 2019; Ferrer-Garcia et al., 2018; Keizer et al., 2014; 2016; Lin et al., 2021; Liu et al., 2022; Malighetti et al., 2021; Neyret et al., 2020; Piryankova et al., 2014; Porras-Garcia et al., 2019; 2021; Preston & Ehrsson, 2014; 2018; Scarpina et al., 2019; Serino et al., 2016; 2017; 2020; Tambone et al., 2021; Themelis et al., 2021; Wolf et al., 2021). Of these 20, five also reported negative modifications (i.e., a more negative rating on a cognitive-affective scale or increased body size estimation/misestimation), generally involving medium or large-sized effects (Ferrer-Garcia et al., 2018; Lin et al., 2021; Preston & Ehrsson, 2018; Tambone et al., 2021; Themelis et al., 2021). Of the remaining four studies, three observed medium or large-sized negative modifications alone (Ferrer-Garcia et al., 2017; Normand et al., 2011; Porras-Garcia et al., 2020), and one observed no significant change (with a small non-discernible effect; Provenzano et al., 2019).

**2a. Clinical conditions and symptoms.** Of the 24 studies, eight assessed a clinical ED sample, of which six (75.0%) observed improvement. Specifically, of the seven examining anorexia exclusively, five generally reported medium or large-sized improvements (Keizer et al., 2014; 2016; Malighetti et al., 2021; Porras-Garcia et al., 2021; Serino et al., 2017), although one reported medium to large-sized negative shifts in BID (Porras-Garcia et al., 2020), and the remaining study observed no significant change in BID (with a small-sized non-discernible effect; Provenzano et al., 2019). One study examined a mixed ED group and reported small or medium-sized improvements (Carey & Preston, 2019). Five of these eight studies included healthy controls: two also observed improvements in BID in healthy controls, although to a lesser extent (generally, small to medium-sized effects) than in the clinical samples (Carey & Preston, 2019; Keizer et al., 2016).

The remaining 16 studies assessed a community-based sample alone. Fourteen (87.50%) reported improvements in BID post-embodiment (generally involving medium to large-sized effects; Ferrer-Garcia et al., 2018; Lin et al., 2021; Liu et al., 2022; Neyret et al., 2020; Piryankova et al., 2014; Porras-Garcia et al., 2019; Preston & Ehrsson, 2014; 2018; Scarpina et al., 2019; Serino et al., 2016; 2020; Tambone et al., 2021; Themelis et al., 2021; Wolf et al., 2021). The remaining two studies reported a negative shift alone (medium to large-sized effects; Ferrer-Garcia et al., 2017; Normand et al., 2011). In three of 16 studies, greater positive (Preston & Ehrsson, 2014; 2018) and/or negative (Ferrer-Garcia et al., 2017; Preston & Ehrsson, 2018) changes were observed among those with higher (relative to lower) BID (e.g., body dissatisfaction), with medium or large-sized effects.

**2b. Embodiment illusion paradigm.** Of the 24 studies, 22 induced a VR-based full-body illusion: 18 (81.80%) found medium or large-sized improvements in BID across perceptual and/or cognitive-affective dimensions (Ferrer-Garcia et al., 2018; Keizer et al., 2016; Lin et al., 2021, Liu et al., 2022; Malighetti et al., 2021; Neyret et al., 2020; Piryankova et al., 2014; Porras-Garcia et al., 2019; 2021; Preston & Ehrsson, 2014; 2018; Scarpina et al., 2019; Serino et al., 2016; 2017; 2020; Tambone et al., 2021; Themelis et al., 2021; Wolf et al., 2021); and four observed medium to large-sized negative modifications alone across both dimensions (Ferrer-Garcia et al., 2017; Normand et al., 2011; Porras-Garcia et al., 2020) or a non-discernible non-significant small change (Provenzano et al., 2019). The remaining two studies used a real-world RHI and found small to large-sized improvements in perceptual BID (Carey & Preston., 2019; Keizer et al., 2014).

**2c. Body part(s) embodied**. Of the 20 studies that found improvements in BID, nine stimulated the abdomen alone, generally involving medium to large-sized improvements (Keizer et al., 2016; Neyret et al., 2020; Preston & Ehrsson, 2014; 2018; Scarpina et al., 2019; Serino et al., 2016; 2017; 2020; Tambone et al., 2021). Two stimulated the hand alone and produced small to large-sized improvements (Carey & Preston, 2019; Keizer et al., 2014). The remaining nine studies stimulated multiple regions simultaneously (e.g., hand, arms, legs), generally producing medium or large-sized improvements (Ferrer-Garcia et al., 2018; Lin et al., 2021; Liu et al., 2022; Malighetti et al., 2021; Piryankova et al., 2014; Porras-Garcia et al., 2019; 2021; Themelis et al., 2021; Wolf et al., 2021).

**2d. Size of embodied body(part).** Twenty-two studies examined the effect of the size of the embodiment model/avatar relative to the participant’s real size. Fifteen examined a larger body(part): five (33.33%) observed small to large-sized improvements in perceptual and/or cognitive-affective BID (Keizer et al., 2016; Lin et al., 2021; Malighetti et al., 2021; Porras-Garcia et al., 2021; Serino et al., 2020); seven observed medium to large-sized negative modifications alone across both BID dimensions (Ferrer-Garcia et al., 2017; 2018; Normand et al., 2011; Piryankova et al., 2014; Porras-Garcia et al., 2019; Preston & Ehrsson, 2018; Tambone et al., 2021); and three observed no significant change (effects revealed small to large negative modifications in perceptual and cognitive-affective BID; Preston & Ehrsson, 2014; Provenzano et al., 2019; Themelis et al., 2021). Eleven studies examined a smaller body(part): ten (90.91%) observed medium to large-sized improvements across both BID dimensions (Lin et al., 2021; Liu et al., 2022; Neyret et al., 2020; Piryankova et al., 2014; Preston & Ehrsson, 2014; 2018; Scarpina et al., 2019; Serino et al., 2016; 2020; Tambone et al., 2021), although one study also observed a small negative modification in cognitive-affective BID (Tambone et al., 2021). The remaining study observed a small non-discernible non-significant change in BID (Provenzano et al., 2019). Twelve studies examined a same-sized body(part): eight (66.67%) observed medium to large-sized improvements across both BID dimensions (Carey & Preston, 2019; Ferrer-Garcia et al., 2018; Keizer et al., 2014; 2016; Neyret et al., 2020; Porras-Garcia et al., 2021; Serino et al., 2017; Wolf et al., 2021); two observed large-sized negative shifts in cognitive-affective BID (Malighetti et al., 2021; Porras-Garcia et al., 2020); and two observed non-significant small-sized improvements (Porras Garcia et al., 2019) or non-discernible small-sized changes (Provenzano et al., 2019).

**2e. Synchrony of stimulation**. Of the 20 studies that found improvements in BID, only 11 compared synchronous versus asynchronous stimulation conditions: six (54.55%) found medium to large-sized improvements in BID for synchronous relative to asynchronous conditions (Carey & Preston, 2019; Neyret et al., 2020; Porras-Garcia et al., 2019; Preston & Ehrsson, 2018; Scarpina et al., 2019; Serino et al., 2016), however, three also noted at least one positive effect (small to large-sized) irrespective of synchrony (Carey & Preston, 2019; Porras-Garcia et al., 2019; Preston & Ehrsson, 2018). The remaining five studies found no significant effect of synchrony on observed improvements (calculable effects were generally negligible; Keizer et al., 2014; 2016; Piryankova et al., 2014; Serino et al., 2017) or failed to report the effect (Preston & Ehrsson, 2014).

### Quality appraisal and risk of bias assessment

As Table 2 shows, the quality evaluation found that most studies were deemed fair or good (46.88% versus 40.62%), fewer were poor (12.50%), and none were excellent. The most commonly met criteria were the use of appropriate test statistics (100% of studies), exact probability values (84.38% of studies), and clear descriptions of results (81.25% of studies) and interventions of interest (78.13% of studies). Potential sources of bias included low statistical power (or failure to conduct a power analysis; 90.63%), non-representative samples (81.25% of studies), poor control of confounding variables (75.00% of studies), and non-psychometrically validated and reliable main outcome measures (53.13% of studies).

## Discussion

The current systematic review addressed two primary questions. First, whether susceptibility to embodiment illusions differed as a function of varying levels of BID. Second, whether improvements occurred in BID after experiencing embodiment illusions. We also explored whether factors (e.g., ED or BDD diagnosis, body part targeted) impacted embodiment susceptibility and improvements. As predicted, susceptibility to embodiment illusions was greater in those with high (relative to low/no) BID, and improvements in BID generally occurred after experiencing embodiment illusions. Both findings appeared to be influenced by various factors.

### Susceptibility to embodiment illusions as a function of BID

Researchers have increasingly investigated susceptibility to embodiment illusions as a function of BID. However, only one prior review (Crespi & Dinsdale, 2019) has examined susceptibility to embodiment illusions across a range of mental disorders and conditions: observing higher susceptibility in 71% of the seven ED/BDD-based studies. This prior review was non-systematic and focused solely on the RHI. The current review, drawing from an extended body of research involving broader embodiment illusion paradigms, similarly revealed that 64% of studies reviewed reported significantly greater susceptibility among those with high (relative to low/no) BID. The effects were generally medium to large-sized, suggesting that the current findings have practical significance.

These findings align with the notion that higher levels of BID may be related to more malleable perceptual body representation, possibly due to fundamental deficits involving multisensory integration: namely, external visual input receiving more weight than either tactile and/or proprioceptive input (e.g., Eshkevari et al., 2012; Gaudio et al., 2014). Deficits in interoceptive processing have also been positively linked to embodiment susceptibility (Bekrater-Bodmann et al., 2020) and consistently observed in BID populations (for a review see Jenkinson et al., 2020). According to predictive coding accounts of EDs (Barca & Pezzulo, 2020), BID-related disorders may involve bodily-self-perception that is less dependent on incoming sensory input and more reliant on higher-level predictions (based on prior beliefs). Importantly, this interpretation may offer valuable insight into the aetiology of these disorders because prior beliefs are not just important for perceptual inference but also action selection and control (goal-directed behaviour). For instance, when considering EDs, reliance on prior beliefs about which body size is to be pursued (e.g., a very thin body; Barca & Pezzulo, 2020) over internal bodily input (e.g., hunger cues) may contribute to the maintenance of the disorder by increasing maladaptive affect and cognitions (e.g., prioritising visual body-related feedback over hunger cues). This may also pose challenges to forming and maintaining an accurate bodily representation (Barca & Pezzulo, 2020).

This account could be extended to Allocentric Lock Theory (Riva & Gaudio, 2018). This neuroscientific framework posits that individuals with EDs hold a ‘locked’ or unchanging distorted body image because they are unable to update their ‘remembered’ body (i.e., an allocentric representation that they constructed long ago) with perceptual information from their ‘experienced’ body (i.e., an egocentric representation). The minority of inconsistent findings (i.e., null between-group differences or reduced susceptibility in the high BID group) may be attributable to methodological issues, as outlined below.

### Experiencing embodiment illusions as a means to improve BID

Only one prior review which examined 13 VR-based studies involving full-body illusions has considered whether embodiment illusions can impact BID (Turbyne et al., 2021); improvements were reported in 69% of studies. Using an extended number of studies (*n* = 24) involving both VR and real-world approaches, we revealed that 83% of studies observed significant, generally medium to large-sized, improvements in BID post-embodiment; supporting the practical and clinical utility of embodiment illusions when improving BID. Current findings, and those of others (e.g., Keizer et al., 2014; Turbyne et al., 2021), are compatible with the idea of utilising fluctuating perceptual and cognitive-affective body representations (as a result of disturbances in multisensory integration) in individuals with BID (Espeset et al., 2012); specifically, BID appears to be able to improve via the integration of an external body(part) into one’s mental bodily-representation.

### Additional factors impacting susceptibility and improvement

#### Clinical condition and symptoms

From a clinical perspective, EDs and BDD have heterogeneous psychopathologies (APA, 2013). However, no prior embodiment illusion research has compared susceptibility or improvement across these disorders (including their sub-types). Only one study within the current review assessed BDD (inhibiting comparison; Kaplan et al., 2014), whilst ED-based studies examined a mixed group or anorexia nervosa exclusively. Although this precludes comparison across ED-subtypes, tentatively, effect sizes suggest that the level of susceptibility is more pronounced in anorexia nervosa (e.g., Lavenne-Collot et al., 2022) than in mixed ED (e.g., Eshkevari et al., 2014) studies. Theoretically, this is unsurprising, given that distorted bodily perception is required to diagnose anorexia nervosa, but not for bulimia nervosa, binge eating disorder, and other specified feeding and eating disorder (APA, 2013). Though neuroscientific evidence directly comparing these disorders is lacking, anorexia nervosa consistently shows alterations in sensory (e.g., vision, touch; Engel & Keizer, 2017) and multisensory (Brizzi et al., 2023) processing. Therefore, disturbances in multisensory integration may be more pronounced in anorexia nervosa than in other EDs, which may ultimately underpin their generally severely disturbed bodily perception (Gaudio et al., 2014). Future researchers should employ sufficiently powered ED samples containing homogenous subtypes to improve understanding of distinct or shared aetiologies and potentially enhance the effectiveness and specificity of prevention and intervention programs. Moreover, future researchers could assess where different manifestations of BID disorders lie on a potential ‘spectrum of susceptibility’; current findings suggest that anorexia nervosa may be more susceptible than other EDs. When considering community-based populations, susceptibility was also positively correlated with core ED-related symptoms including binge eating/purging (e.g., Metral et al., 2017); suggesting that a more malleable bodily perception may also underpin heightened susceptibility even in individuals with non- and sub-clinical BID.

Regarding improvements in BID, no studies assessed whether this varied across ED subtypes or employed a BDD sample. This should be a focus for future researchers to ensure that embodiment illusions are utilised among those who stand to gain the most benefit. Most clinical ED studies examined anorexia nervosa exclusively, the majority (71%) of which observed medium to large-sized improvements in BID post-embodiment, including reduced overestimation (i.e., of the hand, abdomen, shoulders, and hips), body dissatisfaction, drive for thinness, and fear of gaining weight (e.g., Keizer et al., 2016; Serino et al., 2017). This suggests that the perceptual experiences of body size in anorexia nervosa, although severely disturbed and notoriously difficult to change in a clinical setting (Gaudio et al., 2014), can be improved post-embodiment for salient (e.g., abdomen, hips) and non-salient (e.g., hands, shoulders) regions. Persistent cognitive-affective disturbances were also improved. Current findings also provide preliminary support for embodiment to improve BID among a mixed ED group (Carey & Preston, 2019). Improvements, though more pronounced in clinical populations, were also observed in healthy controls (e.g., Keizer et al., 2016) and community-based samples (e.g., Ferrer-Garcia et al., 2018). These results suggest that embodiment illusions may not only help to improve ED symptoms in clinical populations but mitigate ED risk in healthy populations.

#### Embodiment illusion method

No prior embodiment illusion reviews involving BID populations have directly compared susceptibility or improvements across VR and real-world paradigms. Nonetheless, regarding susceptibility, most studies (89%) using real-world paradigms (e.g., RHIs, enfacement illusions; e.g., Eshkevari et al., 2012) observed greater susceptibility among those with high (relative to low/no) BID, generally involving medium to large-sized effects; whilst fewer VR-based (e.g., full-body illusion) studies (20%) mirrored these findings and produced small to large effects. Given neuroscientific evidence that the brain is more responsive to real-world versus synthetic stimuli typically used in VR situations (Kober et al., 2022), this distinction between methodologies is important in suggesting that real-world paradigms may be a more sensitive means for uncovering differential embodiment susceptibility, and ultimately, the mechanisms underpinning BID.

Curiously, despite the small number of real-world investigations, improvements in BID across studies appeared larger and more pronounced when using VR versus real-world paradigms. Although real-world methods may elicit stronger neural responses, we speculate that VR-based highly customisable exposures to different body dimensions (and appearances) on an individual-to-individual basis (Turbyne et al., 2021) may be a more important factor when inducing improvements. Future researchers should better determine why different methods may influence susceptibility and improvements in apparently contradictory ways.

#### Body part(s) embodied

Existing research has rarely compared the body part(s) embodied in BID populations. Current findings suggest that individuals with high (relative to low/no) BID experienced greater embodiment when stimulation involves the hand (78% of hand-based studies) or the face alone (100% of face-based studies), generally producing medium to large-sized effects; not the abdomen (33% of abdomen-based studies) or an array of body parts (e.g., abdomen, legs, and arms; 0% of studies), producing small to large-sized effects. As individuals with high BID experience deficits in sensory processing (e.g., Engel & Keizer, 2017), differences in susceptibility may be more readily uncovered via illusions stimulating bodily regions of higher (e.g., face, hands) relative to lower (e.g., arms, legs) sensory receptor density in the brain (Corniani & Saal, 2020).

Again, despite findings involving susceptibility, improvements in BID were more pronounced when stimulating the abdomen, followed by multiple regions (e.g., arms, legs, abdomen), and lastly, the hands; though, effect sizes were similar (medium to large) for abdomen and multiple-region focused studies and small to large for hand-based studies. To explain these contradictory findings, saliency may amplify improvements regardless of susceptibility, given that the abdomen holds more cognitive-affective salience in BID populations than the hands (note, the face—a very salient body part—has not been investigated). However, intersectionality between body part and method may be at play here: as a possible confound, all studies using real-world paradigms (note, there were only two for Aim 2) stimulated the hand, whilst VR-based studies stimulated other (including multiple) body parts. Well-designed future studies should directly test each factor to disentangle these contradictory findings involving both method and body part and more broadly, assess whether the relationship between illusion strength and improvement is linear or non-linear.

#### Size of the embodied body (part)

No studies have investigated or reported whether the size of the model/avatar influences susceptibility. This could be important as the broader embodiment illusion literature notes that closely matching the participant and model’s size increases illusion strength (e.g., Kim et al., 2020); however, high BID populations typically overestimate their body size more than low BID populations. Therefore, future researchers should consider controlling for induction of the illusion with models matched to one’s real versus perceived body size as between-group differences in susceptibility may be more pronounced (larger) when controlling for this possible confound.

There is growing acknowledgement that embodied body size may be an influential factor when improving BID. One prior review (Turbyne et al., 2021) reported greater improvements in BID post-embodiment of a smaller (compared to larger) body; however, their findings were limited to VR-based full-body illusions and neglected same-sized bodies. Our findings revealed that improvements in BID were larger and more pronounced post-embodiment of a smaller body (part) relative to one’s actual size (91% of studies; generally involving medium to large-sized effects), whilst fewer studies mirroring these findings for a body(part) of the same size (67% of studies; medium to large effects) or larger (33% of studies; small to large effects). The perceived socio-cultural desirability of the embodied body size may explain these findings. Embodying a ‘thin ideal’ (smaller) body may lead individuals to absorb the socio-culturally desirable attributes of the thin ideal (e.g., Neyret et al., 2020), thus improving affective-cognitive BID to a greater extent than embodying a same-sized or larger body. Although seemingly counter-intuitive, improvements involving BID post-embodiment of a same-sized body(part), though not markedly different from their everyday life, may also be explained by a general overestimation of the actual body size. Given that individuals of low and healthy weight (the majority of the included samples) tend to overestimate their actual body size (Cornelissen et al., 2012), most participants may have perceived the same-sized body as smaller than themselves leading to improved BID.

#### Synchrony of stimulation

It remains unclear whether the effects of temporal synchrony manifest differently in BID populations. Sixty percent of all studies included both synchronous and asynchronous stimulation conditions. Of those that compared both conditions, most populations still experienced embodiment and improvements post-asynchronous stimulation (small to large-sized improvements), though, usually to a lesser extent than post-synchronous stimulation (generally medium to large improvements). Current findings and those by Turbyne et al. (2021) support notions that synchronous stimulation may be more reliable and profound (Botvinick & Cohen, 1998). Interestingly, however, 80% of studies that assessed the effect of temporal synchrony observed greater susceptibility among individuals with high (relative to low/no) BID irrespective of synchrony (generally medium to large-sized effects). As outlined above, individuals with high BID may place greater emphasis on or are more sensitive to visual input during multisensory processing (e.g., Keizer et al., 2014). Thus, in most cases, observing a physically similar, fake body part from a first-person perspective receive stimulation in a congruent position/location, was sufficient to induce the illusion, despite the perceptual alienation of temporally incongruent stimulation. Another possible explanation is that individuals with high (relative to low/no) BID have an increased temporal binding window (Prikken et al., 2019): i.e., their time to perceive two stimuli from different modalities as having a similar source may be larger, leading to a blurry demarcation between synchrony conditions, and a stronger illusion. Ultimately, this may lead to a more malleable body representation.

### Limitations and strengths of the included studies

Confidence in the validity and reliability of our findings should be considered against the limitations and strengths of the included studies. The qualitative assessment revealed that most studies were deemed fair or good, none were excellent, and a small portion was poor. Most studies described the embodiment procedure with a replicable level of detail, clearly reported their results, and randomised their stimulation conditions to reduce the potential confound of order effects. These practices are recommended in future research. Although consistent findings were observed across lower and higher-quality studies (e.g., Eshkevari et al., 2014; Mussap & Salton, 2006), several unaddressed limitations warrant attention.

First, measures of embodiment were substantially heterogeneous, lacked psychometric data, and were generally narrow in scope. Over 40% of studies failed to measure (e.g., Mussap & Salton, 2006) or independently assess (Metral et al., 2017) both subjective and objective embodiment. This is limiting as they offer contrasting and complementary sources of information regarding the mechanisms underlying self-perception (for review see Braun et al., 2018). For subjective embodiment, most researchers created modified versions of two commonly used RHI questionnaires (Botvinick & Cohen, 1998; Longo et al., 2008), which were not validated psychometrically and varied in the number of items, number of sub-scales (ownership, agency, and/or self-location), and scoring (totals, individual sub-scales, or individual items). Body-part size estimation, among the most frequently used measures of objective embodiment (e.g., Keizer et al., 2016), also varied considerably, including width estimates using a laser beam (e.g., Serino et al., 2020) or circumference estimates using string (e.g., Serino et al., 2016). Moreover, these typically failed to capture structural aspects (e.g., the shape of the abdomen), and again, lacked psychometric data. Increased efforts are needed to utilise valid, reliable, and consistent measures of subjective embodiment, such as Romano et al.’s (2021) RHI questionnaire or Gonzalez-Franco and Peck’s (2018) proposed (though not yet validated) standardised embodiment questionnaire. More advanced and controlled body-part measures also exist, such as the Body Image Assessment Software (Ferrer-García & Gutiérrez-Maldonado, 2008) and VR-based methods whereby participants adjust their body size in VR (Provenzano et al., 2020). Although apparent contradictions in our findings may be attributable to the measures used, the heterogeneity and lack of psychometric data inhibit conclusions.

Considerable heterogeneity regarding the scope and psychometric data of measures of BID used in studies may explain some inconsistent findings. For instance, the only study (Provenzano et al., 2019) where embodiment did not affect BID used a non-validated measure of cognitive-affective BID, whilst most studies that observed an effect employed validated measures of cognitive-affective BID or both perceptual and cognitive-affective measures. Future research should utilise a more standardised and advanced battery of valid and reliable measures that span both perceptual and cognitive-affective dimensions of BID as these may map onto objective and subjective measures, respectively.

Second, the inadequate control of confounds may explain inconsistent findings regarding susceptibility. Almost all studies that found non-significant between-group differences or observed greater susceptibility in healthy controls than the high BID groups failed to screen for pre-existing BID among the healthy controls (e.g., Kaplan et al., 2014); the remaining study observed initial body size overestimations in the healthy controls (Carey & Preston, 2019). Thus, high BID may have existed within each healthy control group, reducing the ability to detect between-group differences. Comparatively, all studies that supported the hypothesis explicitly screened for BID-related psychopathology in both groups.

Finally, most participants were adult women from European countries, which is limiting given that significant differences in BID psychopathology exist between men and women (e.g., more muscularity-orientated presentations in men; Murray et al., 2018) and adolescents and adults (e.g., difficulty eliciting change among adults; Porras-Garcia et al., 2021). Non-European Union countries (e.g., the United Kingdom and the United States) and Asia were also neglected which may limit the generalisability of findings given geographical differences involving BID (Frederick et al., 2016). Future researchers should investigate whether and how demographic differences manifest in the current research questions.

### Limitations and strengths of the current review

As discussed, the most notable limitation of the current review was considerable heterogeneity across studies in the assessed populations (e.g., mixed ED group, anorexia nervosa), interventions (e.g., real-world versus VR, body part[s] targeted), and outcome measures. This limited our ability to draw comparisons and conclusions involving a meta-analysis, and/or to construct valid funnel plots indicating potential publication biases: impairing attempts to fully understand the findings including the potential intersectionality between various factors raised.

Second, the current review omitted psychiatric disorders and developmental conditions that show high comorbidity with EDs and BDD, including anxiety and depression (Keski-Rahkonen & Mustelin, 2016), and autism spectrum disorder (Huke et al., 2013). Crespi and Dinsdale’s (2019) review indicated that, relative to healthy controls, individuals with EDs and autism spectrum disorder show increased and reduced susceptibility, respectively. Hence if individuals in the current review had autism spectrum disorder-related ailments, this may underpin some of our observed contradictory findings. Future reviewers should examine studies that have assessed comorbid disorders and conditions in BDD/ED samples (e.g., Kaplan et al., 2014).

Finally, we conceptualised *improvement* in perceptual BID as (i) reduced body size *misestimation* relative to baseline (e.g. Keizer et al., 2014), or (ii) reduced body size *estimation* relative to baseline when data on baseline misestimation was not available (e.g., Wolf et al., 2021). The latter may be problematic as without baseline data, it is speculative that the assessed sample overestimated their body size at baseline (thus, reductions might have reflected BID increases). However, given extensive research demonstrating that BID populations (Hosseini & Padhy, 2019) and those of low and healthy weight (the majority of the included samples; Cornelissen et al., 2012) overestimate their body size, we deemed it valid to infer that the included samples generally overestimated. Nonetheless, future researchers should be more rigorous and assess *misestimation*.

Notwithstanding these limitations, the current review had several strengths. First, including a large number of studies across various embodiment approaches allowed a more exhaustive assessment. Second, including unpublished studies reduced the potential impact of publication bias if only peer-reviewed publications were included (Borenstein et al., 2021). Finally, our quality appraisal and risk of bias assessment ensured that studies of lower quality were interpreted tentatively, and allowed us to provide specific methodological recommendations for future researchers.

### Conclusions, implications, and directions for future research

The heterogeneity and limitations in methodology across studies, as well as the lack of theoretical underpinnings in the field, ultimately warrant caution when making scientific advances on the major components of BID. Nonetheless, multiple take-home messages emerged from our review. First, insights from embodiment illusion experiments may enhance our understanding of the multisensory integration basis of perceptual distortions within BID populations, and ultimately, enhance interventions. Indeed, greater susceptibility was consistently observed in high BID populations and may indicate a more malleable bodily representation due to underlying disturbances in multisensory integration and increased weighting of higher-order predictions over lower-level sensory inputs.

Second, embodiment illusions also provided a means to manipulate and improve the malleable body representation of individuals with BID. This warrants continued attempts to incorporate embodiment illusions within clinical settings as a complementary intervention to best-practice treatments (e.g., CBT) to target more holistic dimensions of BID, and in prevention and early intervention efforts targeted towards BID-related disorders. Given the aforementioned issues associated with CBT (Atwood & Friedman, 2020), focusing on perceptual aspects via embodiment may provide a means to increase the effectiveness and completion of these current interventions.

Third, new therapeutic interventions targeting BID may also utilise insights gained from research on susceptibility to embodiment illusions. For instance, sensory and multisensory training/therapies (e.g., physical therapy, mindfulness-based interventions; Weng et al., 2021) and wearable technologies (e.g., brain/body stimulation devices; Riva et al., 2017) could increase tactile, proprioceptive, and interoceptive awareness to restore the balance between visual and tactile/internal bodily input during multisensory integration (Eshkevari et al., 2014). Ultimately, this may help to form and maintain a more realistic self-representation and thus, improve BID.

Fourth, our exploration of the various influencing factors revealed that susceptibility consistently varied as a function of BID, especially when assessing anorexia nervosa samples, using a real-world embodiment paradigm, and the hand or face to induce the illusion, irrespective of stimulation synchrony. Additionally, improvements in BID were greater when utilising VR-based embodiment illusions, body parts of high cognitive-affective salience, a smaller-sized model/avatar, and synchronous stimulation. These findings may inform future attempts in both contexts to improve the efficacy of future research, prevention, and intervention approaches in BID populations.

Additional future directions warrant attention. First, only three included studies employed longitudinal designs (e.g., Porras-Garcia et al., 2021). Whilst all found enduring improvements in BID post-embodiment, there was considerable heterogeneity in the assessed periods (ranging from a few hours to 3 months). Further research into the longevity of any improvement is required before these methods can be considered in long-term strategies targeting BID. Second, there is a clear gap in embodiment illusion research examining BDD. One study assessed susceptibility in a BDD sample, no studies have examined BDD subtypes (e.g., muscle dysmorphia), and measures of improvement typically neglect BDD-specific constructs. This research would improve understanding of distinct or shared aetiologies across ED and BDD sub-types and thus, the efficacy of embodiment illusion-based interventions. Third, researchers should consider exploring neurobiological underpinnings of susceptibility to embodiment and post-embodiment improvements in BID via neuroscience technologies (e.g., fMRI), to enrich our understanding of whether and how the brain is involved. Current research has implicated—but not directly tested—(i) the insula in susceptibility to the RHI (e.g., Crespi & Dinsdale, 2019) and (ii) interactions between the insula, cingulate cortex, and parietal cortex in the effects of illusory obesity on body dissatisfaction (Preston & Ehrsson, 2016). Finally, there is a risk that ‘passively’ generated change (i.e., independent of the subject’s will)—as arguably observed in embodiment illusions—could produce mostly immediate but then ineffective effects in terms of care and treatment. Future researchers should evaluate whether and by which means embodiment illusions are efficacious in bringing about constant change in an individual who, based on an active process of taking a stand in their symptoms, has found no better way of adaption than to develop their own disorder. Addressing such avenues for future exploration would help to facilitate the successful use of embodiment illusions among researchers and practitioners working with BID populations.

**Table 1.**
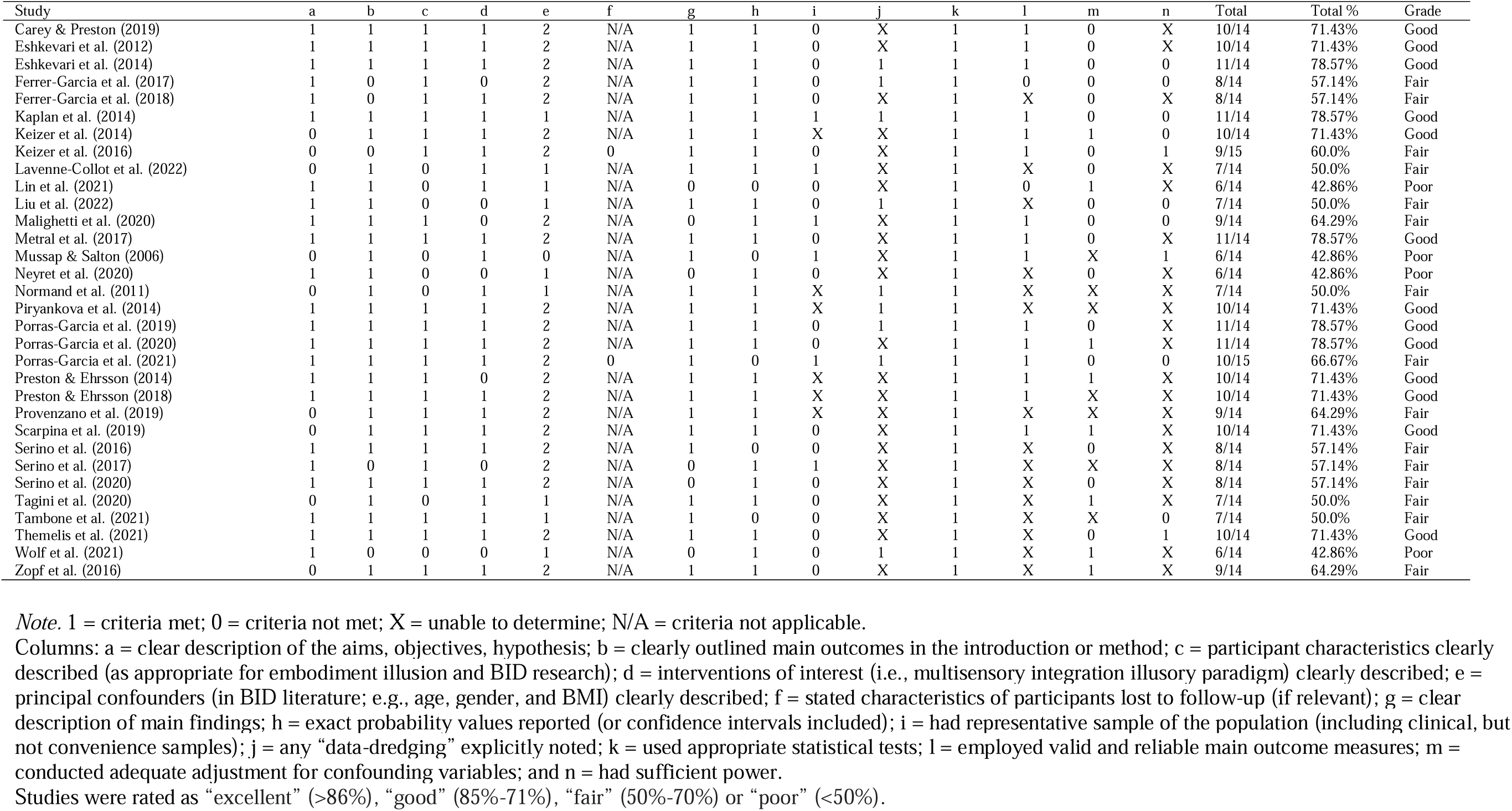
Downs and Black (1998) Checklist for Methodological Quality, Adapted to Evaluate Studies Identified in a Systematic Review Assessing Embodiment Illusions in Relation to Body Image Disturbance.

## Data Availability

All data produced in the present study are available upon reasonable request to the authors

