## Appendices for "Your body, my experience: A systematic review of embodiment illusions as a function of and method to improve body image disturbance"

| **Appendix A. Table 1**  *Common subjective and objective measures of various embodiment illusions* | | |
| --- | --- | --- |
| Illusion | Measures | Description |
| Rubber Hand | Subjective |  |
|  | Embodiment questionnaires (Eshkevari et al., 2012; 2014; Keizer et al., 2014; Mussap & Salton, 2006; Zopf et al., 2016) | - Typically contains between 6 and 12 items scored on Likert scales assessing the intensity of the illusory experience; namely: self-location (i.e., spatial location of one’s own hand), agency (i.e., control) over the rubber hand, and/or ownership (i.e., self-attribution of the rubber hand). - Higher scores indicate greater embodiment. |
|  | Objective |  |
|  | Proprioceptive drift (Eshkevari et al., 2012; 2014; Keizer et al., 2014) | - Participants estimate the perceived position of their unseen finger/hand relative to where the artificial finger/hand is, before and after the illusion. - Calculated as a difference score before and after the illusion. Positive values indicate a larger bias in proprioceptive judgment towards the rubber hand’s location (i.e., stronger embodiment). |
|  | Hand size estimation (Keizer et al., 2014) | - Participants estimate the width and/or length of their hand/wrist and the rubber hand/wrist before and after the illusion. Specifically, participants use a calliper to indicate when the hand fits exactly in between two pointers while the experimenter moves them away from and towards each other. - Calculated as a difference score before and after the illusion. Positive values indicate a change in size estimation after the illusion (i.e., stronger embodiment). |
|  | Reaching task (Zopf et al., 2016) | - An action-orientated measure of body location perception after each illusion induction (e.g., participants use their unseen hand to reach towards visual targets on a screen) - Calculated as ‘reach endpoint errors’ (i.e., the difference between the participant’s movement endpoints and actual target locations). Greater errors indicate greater embodiment. |
|  | Onset latency (Metral et al., 2017), | - Experimenters records delay (onset latency) between beginning of trial and moment the participant reports having felt the illusion (in seconds). Lower onset latency scores indicate greater embodiment. |
| Full Body | Subjective |  |
|  | Embodiment quesionnaires (Keizer et al., 2016; Piryankova et al., 2014; Serino et al., 2019; Provenzano et al., 2019; Tambone et al., 2021; Wolf et al., 2021) | - Contains between 2 and 34 items scored on Likert scales typically assessing experiences of self-location (i.e., spatial location of one’s own body or body part), agency (i.e., control) over the body(part), and/or ownership (i.e., self-attribution of the body or body part). Additional experiences, though less commonly assessed, may include referral of touch (i.e., feeling of being touched by a virtual ball), intensity of touch, identification with avatar, and out-of-body experiences. - Higher scores indicate greater embodiment. |
|  | Visual analogue scales (Porras-Garcia et al., 2019; 2020; 2021) | - Participants estimate the intensity of the illusion (typically ownership) on a scale ranging frm 0 to 100. Higher scores indicate greater embodiment. |
|  | Objective |  |
|  | Body size estimation (Keizer et al., 2016; Serino et al., 2016; 2017; 2019; Scarpina et al., 2019; Tambone et al., 2021) | - Before and after illusion induction, participants estimate the width of different body parts (e.g., shoulders, abdomen, and hips). E.g., by (i) placing adhesive markers on a wall, (ii) marking on a blackboard, (iii) using a laser beam, or (iv) wearing a blindfold, outstretching their arms and adapting the width between their palms. Participants may also estimate the circumference of body parts using string/rope placed on floor. - Misestimation of body size post-embodiment (relative to pre-embodiment) in direction of model’s/avatar’s size reflects greater embodiment. |
|  | Affordance estimation (Piryankova et al., 2014) | - Participants estimate width of their body (parts) (e.g., shoulders). E.g., by adjusting the distance between two poles that cast a shadow on the floor. Scored the same as body size estimation. |
|  | Skin conductance response (Preston & Ehrsson, 2014) | - Experimenters typically attack the participant’s embodied body part with a threatening object (e.g. knife or hammer) to heighten skin conductance as if the real body was being threatened. - Higher scores indicate greater embodiment. |
|  | Body temperature (Provenzano et al., 2019) | - Often measured via an infrared thermometer placed on participant (e.g., under armpit) post-illusion. - Based on notions that a disruption in temperature regulation is linked to a disruption in ownership of the real body, a decrease in skin temperature in the participant’s body indicates greater embodiment. |
| Enfacement | Subjective |  |
|  | Enfacement questionnaires (Ma et al., 2016; Estudillo et al., 2017) | - Typically contains between 7 and 12 items scored on Likert scales assessing experiences of perceived ownership and agency (control) over the other’s (virtual or real) face, appearance similarity with the other’s face, and/or referral of touch (i.e., feeling the touch to the other’s face on one’s own). - Higher scores indicate greater enfacement. |
|  | Objective |  |
|  | Self-other discrimination task (Estudillo & Bindemann, 2017) | - Participants typically watch a video of morphed images gradually transitioning from a face that is 100% model (and 0% self) to 100% self (and 0% model) and are required to stop the video when the images appear more like the self than the model (or vice-versa). - Enfacement is believed to have occurred if participants accept a larger percentage of the model’s facial features as their own following interpersonal multisensory stimulation (relative to baseline), indicating that the model’s appearance has been adopted into their self-perception. |

*Note*. Measures included in this table are those most commonly used in the field and do not reflect all measures utilised among the included studies.

**Appendix B. Table 2**

*Database search terms using boolean operators*

| Conjunction | Search Terms (and Conjunctions) | Key Concept |
| --- | --- | --- |
|  | embodiment OR enfacement OR rubber hand illusion OR full body illusion OR (multisensory adj integrat* OR process* OR stimulat* OR illus*) OR (body OR bodily adj illus*) OR (visuo-tactile adj synchron* OR “stimulat*) OR virtual reality | Multisensory Integration Illusory Paradigm |
| AND | (body OR bodily OR image OR weight) adj2 (satisf* OR dissatisf* OR disturb* OR distort* OR anxiety) OR | Body Image |
|  | (face OR facial) adj2 (satisf* OR dissatisf* OR disturb* OR distort* OR anxiety) OR |  |
|  | self-perception OR self-recognition |  |
| AND | exp eating disorders/ OR | Eating Symptomatology |
|  | (eating) adj2 (symptomatology OR psychosymptomatology OR pathology OR psychopathology) OR |  |
|  | ((eating OR appetite) adj (disorder* OR disease* OR illness* OR addiction*)) OR |  |
|  | (anorexi* OR bulimi* OR binge eating OR binge purg* OR purging OR disordered eating OR dietary restr* OR feeding disorder* OR hyperphagia OR pica) OR |  |
|  | eating behaviour* OR eating attitude* OR |  |
|  | weight adj2 (gain* OR los* OR control*) |  |
| AND | body dysmorph* OR muscul* dysmorph* OR muscul* dissatisf* OR muscul* satisf* | Dysmorphic Symptomatology |

*Note.* Adj = adjunct. We included the broader term “virtual reality” since not all studies containing full body illusions mention this term explicitly in the title, abstract, or keywords. Specific key words and free text terms were combined with Boolean operators according to the different terms and rules for each database.

**Appendix C. Table 3**

*Overview of studies investigating embodiment illusions in clinical ED or BDD and community-based samples*

| **Study** | **Aim/s** | **Participants, Group/s, and Country** | **Stimuli/Measures** | **Procedure** | **Key Findings*** |
| --- | --- | --- | --- | --- | --- |
| Carey & Preston (2019) | 1, 2 | *N* = 50 females  ED group (inpatient and outpatient sample):  *n* = 26: 18 (AN), 1 (BN), 2 (BED), 5 (OSFED); *M* _age_ = 23.46 years, *SD* = 5.95; *M* _BMI_ = 19.80, *SD* = 4.39  HC group (university sample):  *n* = 24; *M* _age_ = 19.13 years, *SD* = 1.42; *M* _BMI_ = 20.75, *SD* = 2.30  Country: United Kingdom | 1.Moving RHI paradigm (variant of the classic RHI paradigm involving mechanical-based movement of the participant’s hand connected to a fake hand)  2. Subjective embodiment: ownership, agency (Moving RHI Questionnaire)  3. Objective embodiment: proprioceptive drift; hand size estimation  4. BID: ED psychopathology (EDE-Q) | Moving RHI: sync and async condition (randomised order across participants; 3 x trials per condition). Visuo-motor stimulation (hand movement).  Subjective embodiment measure after each condition. Hand size estimation and proprioceptive drift at baseline and after each condition. | **Subjective embodiment**  Ownership and location:  - Sync > async (*d*’s > 3.10)  - HC = ED (*d*’s = 0.12-0.30; ED > HC)  - Entire sample: positive correlation with implicit body satisfaction, driven by ED psychopathology (*d* = 0.68); no correlation with ED psychopathology (*d* = 0.10-0.12; positive and negative correlation)  **Objective embodiment**  Proprioceptive drift:  - Sync = async (*d* = 0.36; direction not discernible)  - HC = ED (*d* = 0.15; ED > HC)  **Objective embodiment and BID**  Hand size estimation:  - Pre-illusion: ED = HC (although both groups overestimated; *d* _ED_ = 0.65; *d* _HC_ = 0.44)  - Post-illusion: Significant reduction in overestimation in ED group (sync = async) and HC group (sync only; *d* _ED_ = 0.54-0.56; *d* _HC_ = 0.43) |
| Eshkevari et al. (2012) | 1 | *N* = 139 females  ED group (university and community sample):  *n* = 78: AN (*n* = 36), BN (*n* = 22), EDNOS (*n* = 20); *Mdn* _age_ *=* 22.5-27.5 years, *IQR =* 10-18 years; *Mdn* _BMI_ = 16.1-20.9, *IQR =* 2.7-5.5  HC group (university and community sample):  *n* = 61; *Mdn* _age_*=* 24 years*, IQR = 7*; *Mdn* _BMI_ = 21.5, *IQR* = 2.8  Country: United Kingdom | 1. RHI paradigm  2. Objective embodiment: proprioceptive drift  3. Subjective embodiment: ownership, agency, and self-location (EQ) | RHI: sync and async conditions (randomised order across participants; 1 x trial per condition). Visuo-tactile stimulation (hands brushed). Proprioceptive drift measures at baseline. Proprioceptive drift and EQ measures after each condition. | **Subjective embodiment**  Ownership, agency, and self-location:  - ED > HC (*d* = 0.56)  - Sync > async (*d* = 1.95)  - Entire sample: positive correlation with drive for thinness, body dissatisfaction (*d*’s = 0.68-0.70), and depressive symptoms (*d* = 0.54); no correlation with bulimia (*d* = 0.28; positive correlation)  **Objective embodiment**  Proprioceptive drift:  - ED > HC (*d* = 0.37)  - Sync > async (*d* = 0.85) |
| Eshkevari et al. (2014) | 1 | *N =* 167 females  ED group (same as Eshkevari et al., 2012)  REC group (university and community sample): *n* = 28; *Mdn* _age_= 25.5 years, *IQR* = 8  HC group (same as Eshkevari et al., 2012)  Country: United Kingdom | 1. RHI paradigm  2. Subjective embodiment: ownership, agency, and self-location (EQ) | RHI: sync and async conditions (randomised order across participants; 1 x trial per condition). Visuo-tactile stimulation (hands brushed). Proprioceptive drift and EQ measures after each condition. | **Subjective embodiment**  Ownership, agency, and self-location:  - ED and REC > HC; ED = REC (*d* = 0.41)  - Sync > async (*d* = 1.84)  **Objective embodiment**  Proprioceptive drift:  - ED = HC = REC (*d* = 0.79; direction not discernible)  - Sync > async (*d* = 0.72) |
| Ferrer-Garcia et al. (2017) | 2 | *N* = 23 (5 male, 18 female) (university sample): *M* _age_ = 24.91 years, *SD* = 4.78; *M* _BMI_ = 22.18, *SD* = 4.41  Country: Spain | 1.FBI paradigm: via HMD  2. Avatars: (1) same body measurements as participant, (2) 20% larger or (3) 40% larger than participant.  3. Subjective embodiment: ownership (VAS-OI)  4. BID: body anxiety (PASTAS); fear of gaining weight (VAS-fear of gaining weight); trait drive for thinness and body dissatisfaction (EDI) | FBI: sync condition (1 x trial per avatar). Visuo-motor stimulation (arm movement).  PASTAS, VAS-fear of gaining weight, and VAS-OI after exposure to avatar (1) and (3). Only the latter two measures after exposure to avatar (2). | **Subjective embodiment**  Ownership:  - High in all body-size conditions (insufficient data to calculate effect sizes)  **BID**  Body anxiety and fear of gaining weight:  - Post-embodiment 40% larger avatar > real-size avatar (both outcomes) and 20% larger avatar (fear of weight gain only) (fear of weight gain [*d*’s > 1.69]; anxiety [*d* = 1.02])  - Both increased after owning the largest avatar only for individuals with high trait body dissatisfaction (body anxiety: *d* *^=^* 1.00; fear of gaining weight: *d* = 0.64) and drive for thinness (body anxiety: *d* = 1.29; fear of gaining weight: *d* = 0.72) |
| Ferrer-Garcia et al. (2018) | 2 | *N* = 40 females (university sample): *M* _age_ = 22.55 years, *SD* = 4.02; *M* _BMI_ = 21.65, *SD* = 0.40  Country: Spain | 1. FBI paradigm: via HMD  2. Avatars (as described in Ferrer-Garcia et al., 2017)  3. Subjective embodiment: ownership (VAS)  4) BID: body dissatisfaction and body distortion (BIAS-BD) | FBI: sync condition (1 x trial per avatar). Visuo-tactile stimulation (touches on arms, legs, abdomen). BIAS-BD measure at baseline. BIAS-BD and VAS measures after each avatar embodiment. | **Subjective embodiment**  Ownership:  - No difference after embodying different-sized avatars (*d* = 0.31; same-size 1^st^ time > larger > same size 2^nd^ time)  **BID**  Body dissatisfaction and body distortion:  - Post-embodiment of same-size avatar 2^nd^ time < larger avatar (40% = 20%), same-size avatar 1^st^ time, and pre-test (Body distortion; same-size 1 vs. 2 [*d* = 0.84]; larger vs. same-size 2 [*d* = 0.68]. Body dissatisfaction; pre-test vs. same-size 2 [*d* = 1.31]; same-size 1 vs. 2 [*d* = 0.67]; larger vs. same-size 2 [*d* = 1.08])  - All other comparisons non-significant: pre-test > same-size 1 (*d’s* = 0.00-0.40); pre-test > same-size 2 (*d* _body distortion_ = 0.45); pre-test > larger (*d*’s = 0.00) |
| Kaplan et al. (2014) | 1 | *N* = 34 (26 females, 8 males)  BDD group (hospital-based body image clinic sample):  *n* = 17 (13 females); *M* _age_ = 36.41 years, *SD* = 11.27  HC group (university and community sample):  *n* = 17 (13 females); *M* _age_ = 35.41 years, *SD* = 9.73  No BMI information.  Country: Australia | 1. RHI paradigm  2. Subjective embodiment: ownership, agency, location (RHI-Q)  3. Objective embodiment: proprioceptive drift  4. BID: ED symptoms (bulimia, body dissatisfaction, drive for thinness; EDI-3); BDD symptoms (dysmorphic concern; DCQ) | RHI: sync and async conditions (randomised order across participants; 4 x trials [2 x sync, 2 x async]).  Visuo-tactile stimulation (hands stroked). BID measures at baseline. Proprioceptive drift and RHI-Q measures completed after each trial. | **Subjective embodiment**  Illusion index:  - BDD = HC (*d* = 0.59; BDD > HC)  - Sync > async (*d* = 2.04)  - Positive correlation between RHI strength and ED/BDD symptoms; bulimia (*d* = 0.92), body dissatisfaction (*d* = 0.84), drive for thinness (*d* = 0.64), dysmorphic concern (*d* =0.52)  **Objective embodiment**  Proprioceptive Drift:  - Sync = async (*d* = 0.45; sync > async)  - BDD = HC (insufficient data to calculate effect size), though BDD group less affected by stimulation synchrony; BDD: (Sync = async) > baseline; HC: Sync > async and baseline; baseline = async (BID group*synchrony: *d* = 0.70)  - No correlation with ED or BDD symptoms (insufficient data to calculate effect sizes) |
| Keizer et al. (2014) | 1, 2 | *N* = 60 females  AN group (patient sample):  *n* = 30; *M* _age_ = 26.37 years, *SD* = 9.08; *M* _BMI_ = 17.50, *SD* = 2.14  HC group (unspecified sample):  *n* = 30; *M* _age_ = 21.80 years, *SD* = 2.37; *M* _BMI_ = 21.19, *SD* = 2.01  Country: The Netherlands | 1. RHI paradigm  2. Subjective embodiment: ownership, agency and self-location (EQ)  3. Objective embodiment: proprioceptive drift; hand size estimation  4. BID: ED psychopathology (EDI-2) | RHI: sync and async conditions (randomised order across participants; 4 x trials [2 sync, 2 async]). Visuo-tactile stimulation (hands stroked). Hand estimation and proprioceptive drift measures at baseline. Proprioceptive drift, hand estimation, and EQ measures after each trial. | **Subjective embodiment**  Ownership:  - Sync > async (insufficient data to calculate effect size)  - Sync: AN > HC (*d* = 0.58); Async: AN = HC (*d* = 0.32; AN > HC)  Agency and self-location (control):  - AN = HC (*d*’s = 0.37-0.45; AN > HC)  **Objective embodiment**  Proprioceptive drift:  - Sync > async (*d*’s = -1.58- -1.59)  - AN = HC (*d*’s = 0.19-0.40; AN > HC)  **Objective embodiment and BID**  Hand size misestimation:  - AN group: Overestimated own hand width (but not wrist width or hand length) pre-and post-illusion, however, post-illusion misestimation < baseline (sync < baseline [*d* = 3.09]; async < baseline [1.99]; two difference scores did not differ [unable to calculate effect size]) - HC group: sync = async = baseline (insufficient data to calculate effect sizes)  - No correlation between any variables with trait ED psychopathology (*d*’s = 0.02-1.09; AN > HCs) |
| Keizer et al. (2016) | 1, 2 | *N* = 59 females  AN group (ED centre sample):  *n* = 30; *M* _age_ = 22.03 years, *SD* = 3.67; *M* _BMI_ = 18.11, *SD* = 1.68  HC group (university sample):  *n* = 29; *M* _age_ = 21.07 years, *SD* = 2.34; *M* _BMI_ = 20.77, *SD* = 1.48  Subset at follow-up: *n* _AN_ = 9; *n* _HC_ = 26  Country: The Netherlands | 1. FBI paradigm: via HMD  2. Avatar: 25-year-old female, BMI, waist-to-hip ratio, and waist circumference considered healthy  3. Subjective embodiment: ownership, agency, and location (EQ)  4. Objective embodiment and BID: body size estimation task (shoulder, abdomen and hip circumference and width, and height) | FBI: sync and async conditions (order randomised across participants; 1 x trial per condition). Visuo-tactile stimulation (stroking on abdomen). Body size estimates at baseline. Body-size estimates and EQ after experiencing both conditions. Follow-up body size estimation measures 2 hours 45-mins after the FBI. | **Subjective embodiment**  Ownership, agency, and self-location:  - Sync > async (*d* = 1.61)  - AN = HC (*d* = 0.00)  **Objective embodiment and BID**  Body size misestimation:  - Pre-embodiment to post-embodiment: AN group > HC: all body parts (*d*’s = 1.22-1.67), except height (*d* = 0.00)  - AN: Pre-illusion > follow-up for shoulder width, shoulder circumference, and hip circumference (*d*’s = 1.07-1.32), not other body parts (*d*’s = 0.02-0.13; pre- > post-illusion all except shoulder width)  - HC: Pre-illusion > follow-up for height, abdomen and hip circumference (*d*’s 0.47-0.69), not other body parts (*d*’s = 0.12-0.33; pre- > post-illusion all except shoulder width) |
| Lavenne-Collot et al. (2022) | 1 | *N* = 14 (12 females, 2 males); *M* _age_ = 15.6 years  AN group (patient sample): *n* = 7  HC group: *n* = 7  Country: France  No BMI information. | 1. Double mirror paradigm (variant of enfacement illusion: participant observes another’s face through a semi-transparent double-mirror; relative light intensity for each participant changes over time to generate different conditions; *Self* [100% self]; *Other* [0% self]; Self-other face images fully fuse when both lights are on)  2. Objective enfacement: face identification (ID) task (participants verbally state who they recognise most in mirror [Self or Other]) | Two illusion conditions:  (1) Neutral; *Other* and *Self* conditions (randomised order for HCs and AN). Face ID task after each change.  (2) Sensorimotor: Same as part 1, however, AN (not HCs) simultaneously completed five different sensorimotor tasks (e.g., weights on wrists/ankles) | **Objective enfacement**  Face identification:  - Neutral condition: AN > HCs (specifically, AN required a lower portion of self to recognise themselves in the image than HCs, irrespective of the direction of morphing) (*d* = 3.23 [Self-Other condition]; *d* = 1.36] Other-Self condition])  - Sensorimotor condition: AN > HCs (as above) in 4/5 conditions (*d*’s > 1.71 [Self-Other condition]; *d*’s > 1.55 [Other-Self condition]) |
| Lin et al. (2021) | 2 | *N* = 96 (51 females, 45 males) (university sample): *M* _age_ = 21.56 years  Country: Taiwan  No BMI information. | 1. FBI paradigm  2. Avatars: (1) tall and muscular body with a six-pack, (2) tall and normal body, (3) short and muscular body, (4) short and normal body; sex and height matched to participant  3. Subjective embodiment (BOI-Q)  4. BID: self-efficacy for exercise (scale unclear); automatic self-concept (IAT) | FBI: sync condition only (completed series of workouts with one avatar; randomly assigned).  Visuo-motor stimulation (standing core workouts, i.e., knee raise, twist toes touch, side crunch, side leg raise, squat). Both measures administered post-FBI. Follow-up IAT measure 24 h post FBI. | **Subjective embodiment**  - No data provided  **BID**  Immediate and next-day self-efficacy for exercise:  - Normal avatar group > muscular avatar group (*d* = 0.43 [immediate]: *d* = 0.39 [next day])    Self-concept (body image):  - Embodying a muscular avatar increased perceived muscularity (females) perception of normal body shape (males). No effect for normal avatar group. (Group*gender; *d* = 0.42) |
| Liu et al. (2022) | 2 | *N* = 77 females (community sample): *M* _age_ = 21.96 years; *SD* = 3.26  No BMI information.  Country: Germany | 1. Body discontinuity paradigm (variant of the FBI involving visual discontinuity in the avatar [e.g., missing body parts, floating head] to reduce embodiment)  2. Avatar: Female; Age; 18-years: BMI; 20.85 (thin-ideal)  3. Subjective embodiment: ownership, agency, and self-location (EQ)  4. BID: implicit and explicit body image; for both, self-perceived thinness and desire for thinness (RRT) | Randomly assigned to one of two avatar conditions: (1) high embodiment/continuity (showed all body parts); (2) low embodiment/ discontinuity (avatar wrists, ankles and neck invisible).  Body discontinuity paradigm: sync condition only (10 x trials). Visuo-motor simulation (moved boxes and full-body pose). Both measures after final trial. | **Subjective embodiment**  Ownership, agency, and self-location:  - Continuity condition > discontinuity condition (*d* = 1.03)  **BID**  Implicit body image  - Actual (self-perceived thinness): Continuity condition > discontinuity condition (*d* = 1.18)  - Ideal (desire for thinness): No difference between conditions (insufficient data to calculate effect size)  Explicit body image  - Actual (self-perceived thinness): Continuity condition > discontinuity condition (*d* = 0.79)  - Ideal (desire for thinness): No difference between conditions (insufficient data to calculate effect size) |
| Malighetti et al. (2021) | 2 | *N* = 7 females with AN (inpatient sample): *M* _age_ = 17 years, *SD* = 1.87; *M* _BMI_ = 15.95, *SD* = 0.61  Country: Italy | 1. FBI paradigm  2. Avatars: BMI; (1) same as participant (underweight), (2) increased to 17.5, and (3) increased to 18.5 (healthy BMI)  3. Subjective embodiment: ownership, agency, and location (modified EQ)  4. BID: body size estimation task (modified avatar’s BMI to match ideal and perceived body); body dissatisfaction (BSS); body shape preoccupations and concerns (BSQ); body shame (OBCS) | FBI: sync condition only. 4 x sessions; avatar (1) in sessions one and two, avatar (2) in session three, and avatar (3) in session four).  Visuo-motor stimulation (moved arms and legs). VR body size estimation, BSS, BSQ, and OBCS measures pre-and post-embodiment, and EQ post-embodiment. | **Subjective embodiment**  Ownership, agency, and self-location:  - Perceived real body size avatar > desired body size avatar (agency: *d* = 2.45; self-location: *d* = 3.34; ownership: unable to calculate)  **BID**  Body size estimation:  - Improved estimation of desired BMI from sessions 1-4 (i.e., desired a body that was closer to a normal BMI than a pathological size) (insufficient data to calculate effect sizes)  Body dissatisfaction, body shame, and body shape concern:  - Body dissatisfaction reduced from sessions 1-4 (specifically, post-embodiment with healthy BMI avatar < underweight BMI avatar); no change for body shape concern or body shame (insufficient data to calculate effect sizes) |
| Metral et al. (2017) | 1 | *N* = 36 females (university sample): *M* _age_ = 21.30 years, *SD* = 6.36; *M* _BMI_ = 21.45, *SD* = 2.87  Country: France | 1. RHI paradigm  2. Subjective embodiment: ownership, agency, self-location (modified EQ)  3. Objective embodiment: proprioceptive drift; onset latency  4. BID: ED-related traits; drive for thinness, bulimia, body dissatisfaction, ineffectiveness, and maturity fears (EDI-2) | RHI: sync and async conditions (randomised order across participants; 2 x sessions of each condition; 2 x trials per condition per session). Visuo-tactile stimulation (hands stroked). Proprioceptive drift, onset latency and EQ measures before and after, during, and after each trial, respectively. | **Subjective and objective embodiment**  Aggregate of EQ, proprioceptive drift, and onset latency values:  - Positive correlation with ED-related traits: bulimia, ineffectiveness, and maturity fears (*d*’s = 0.82-0.95); no correlation with body dissatisfaction or drive for thinness (*d*’s = 0.04-0.12; positive correlations)  - Sync > async (EQ: *d* = 3.20; proprioceptive drift: *d* = 1.99; onset latency: *d* = 2.64) |
| Mussap & Salton (2006) | 1 | *N* = 128 (70 females, 58 males) (university-dominated sample)  *analysis limited to 101 (right-sided preference, (45% male, 55% female)  No age/BMI information  Country:  Australia | 1. RHI paradigm  2. Subjective embodiment: ownership, agency, location, and global body configuration (RHI-Q)  3. BID: ED-related traits; drive for thinness, bulimia, and body dissatisfaction subscales (EDI-2); unhealthy body development strategies; dietary supplements, steroids, exercise (BCI) | RHI: sync condition only (1 x trial per condition [left or right hand]; randomised order across participants). Visuo-tactile stimulation (hands brushed). All measures after the paradigm. | **Subjective embodiment**  Ownership, agency, self-location, and global body configuration:  - Positive correlation with ED-related traits: bulimic and unhealthy body development (left hand) (*d*’s = 0.52-0.56); no correlation with body dissatisfaction or drive for thinness (*d*’s = 0.06-0.20; positive correlations for left hand; negative correlations for right hand) |
| Neyret et al. (2020) | 2 | *N* = 23 (11 males, 12 females) (university sample): *M* _age_ = 24.8 years, *SD* = 5.64  No BMI information  Country: Spain | 1. FBI paradigm: via HMD  2. Avatars: Three gender-matched avatars, representing the participants (1) ideal body (generally reflected ‘thin ideal’ hourglass shape), (2) real body, and (3) body image.  3. Subjective embodiment: ownership (BOQ)  4. BID: evaluate appearance (body shape, attractiveness) of avatar representing one’s own real and ideal body | FBI: sync (1PP) and async (3PP) conditions (3 x trials per condition, i.e., 1 trial per avatar).  Visuo-motor stimulation (head movements) and visuo-tactile stimulation (vibrations on hands and legs).  Session 1: EDI-2 and BSQ. Session 2 (one week later): paradigm then all measures. | **Subjective embodiment**  Ownership:  - Sync (1PP) > Async (3PP) (irrespective of avatar's body)  **BID**  Body dissatisfaction and drive for thinness:  - Post-embodiment < pre-embodiment (females only) (did not assess effects of avatar size or perspective/synchrony)  Avatar evaluation (index of BID):  - Evaluated real and ideal body (conformed with thin-ideal) as thinner post-embodiment (irrespective of perspective/synchrony) and more attractive (when presented as someone else's [3PP] than when perceived as one's own [i.e., from an embodied 1PP]) (females only)  - Desired own body to resemble real body avatar only (from 3PP only) (no effect of perspective for males)  (Insufficient data to calculate effect sizes) |
| Normand et al. (2011) | 2 | *N* = 22 males (unspecified sample): *M* _age_ = 26.0 years, *SD* = 5.0  No BMI information  Country: Spain | 1. FBI paradigm: via HMD  2. Larger avatar: belly size; 45 centimetres between the avatar’s spine and belly button; rod used to deliver stimulation to belly and estimate belly size  3. Subjective embodiment: ownership, agency, and self-location (modified EQ)  4. Objective embodiment and BID: body (belly) size estimation | FBI: sync and async conditions (randomised order across participants; 1 x trial per condition).  Visuo-tactile stimulation (stomach was poked). Belly size estimates at baseline and post-embodiment, EQ also post-embodiment. | **Subjective embodiment**  Ownership, agency, and self-location:  - Sync > async (insufficient data to calculate effect size)  **Objective embodiment and BID**  Size change (body size overestimation):  - Sync > async (insufficient data to calculate effect size)  - Positive correlation with embodiment strength (irrespective of synchrony; *d* = 0.68) |
| Piryankova et al. (2014) | 2 | *N* = 32 females (unspecified sample): *M* _age_ = 26.0 years; *M* _BMI_ = 22.08, *SD* = 2.95  Country: Germany | 1. FBI paradigm: via HMD  2. Female avatars: width of hips and shoulders adjusted to resemble (1) underweight (BMI of 16), and (2) overweight (BMI of 43); leg, arm- and torso length matched to participant  3. Subjective embodiment: assessing ownership, agency, and self-location (modified EQ)  4. Objective embodiment and BID: body size estimation task for *experienced* body (body they feel at that moment); affordance estimation task | Participants randomly assigned to either avatar. FBI: sync and async conditions (randomised order across participants; 1 x trial per condition).  Visuo-tactile stimulation (stroking on arms and legs). Four trials of affordance and body size estimates before and after embodiment procedure. EQ after both conditions. | **Subjective embodiment**  Ownership, agency, and self-location:  - Sync > async (*d*’s = 1.14-3.52)  **Objective embodiment and BID**  Body size and affordance estimates:  - Both avatar body sizes biased *experienced* body size (*d* = 1.19) and affordances (*d* = 1.01) (*d*’s = 0.00-0.12; direction not discernible) |
| Porras-Garcia et al. (2019) | 2 | *N* = 50 (40 female, 10 male) (university sample): *M* _age_ = 21.8 years, *SD* = 2.55; *M* _BMI_ = 22.50, *SD* = 2.51  Country:  Spain | 1. FBI paradigm: via HMD  2. Avatars: (1) same weight as participant; (2) larger than participant  3. Subjective embodiment: ownership (FBI-VAS)  4. BID: body anxiety (PASTAS); body image distortion and body dissatisfaction (BIAS-BD); fear of gaining weight (VAS) | FBI: sync or async conditions (between-subjects) with both avatars (within-subjects). Visuo-tactile stimulation (touches to arms, legs, and abdomen). BIAS-BD, PASTAS, fear of gaining weight VAS at baseline and all measures including FBI-VAS after FBI. | **Subjective embodiment**  Ownership:  - Sync group > async group (*d* = 0.75)  - Post-larger-size avatar < post-real-size avatar (*d* = 0.50)  **BID**  Body anxiety, body image distortion, body dissatisfaction, and fear of gaining weight:  - Post-embodiment larger avatar > post-same-size avatar and pre-test (body anxiety: *d* = 0.77; body image distortion: *d* = 0.92; body dissatisfaction: *d* = 0.75; fear of gaining weight: *d* = 1.28)  - Baseline = post-embodiment same-size avatar (all scales: *d* = 0.27; generally, baseline > post-embodiment same-size)  - Sync group > async group (body anxiety only; *d* = 0.42). Remaining scales (*d*’s = 0.00-0.47; sync > async) |
| Porras-Garcia et al. (2020) | 1, 2 | *N* = 73 females  AN group (inpatient sample): *n* = 30; 23 (restrictive AN); 7 (purgative AN); *M* _age_ = 17.73 years; *SD* = 4.60; *M* _BMI_ = 17.55, *SD* = 1.07  HC group (university sample): *n* = 43; 25 (low body dissatisfaction), 18 (high body dissatisfaction); *M* _age_ = 21.12 years; *SD* = 1.56; *M* _BMI_ = 21.94, *SD* = 2.53  Country: Spain | 1. FBI paradigm: via HMD  2. Avatar: young female; BMI and silhouette (shoulders, arms, chest, waist, stomach, hip, thighs and legs) matched the participant's real BMI and silhouette  3. Subjective embodiment: ownership (FBI-VAS)  4. BID: body dissatisfaction, drive for thinness (EDI-3); body anxiety (PASTAS); body image disturbance (BIAS); body appreciation (BAS); FGW (VAS) | FBI: sync condition only (1 x trial for each mode of stimulation).  Visuo-tactile stimulation (touches on legs, arms, and stomach) and visuo-motor stimulation (movement). EDI-3, PASTAS, BAS at baseline, then all VASs (FBI, fear of gaining weight, body anxiety) whilst immersed in VR. Body-related attentional bias assessed post-FBI. | **Subjective embodiment**  Ownership:  - AN < HC (low body dissatisfaction only; *d* = 0.77)  - AN: negative correlation with body distortion (*d* = 0.94) and body dissatisfaction (*d* = 0.97); no correlation with remaining BID measures (*d*’s – 0.03-0.75; negative and positive correlations)  - HC (high body dissatisfaction): no correlation with BID (*d*’s = 0.07-0.96; mostly negative correlations); HC (low body dissatisfaction): no correlation with BID (*d*’s = 0.06-0.56; mostly negative correlations)  **BID**  Fear of gaining weight:  - AN > HC (low body dissatisfaction only) post-embodiment (*d* = 1.34)  - HCs: high > low body dissatisfaction post-embodiment (insufficient data to calculate effect size)  Body anxiety:  - AN = HC post-embodiment (*d* = 0.49; AN > HC)  - HCs: low = high body dissatisfaction post-embodiment (insufficient data to calculate effect size)  Body-related attentional bias:  - AN > HC (high and low body dissatisfaction) post-embodiment (all weight-related areas) (*d*’s > 1.21-1.40)  - HCs: low = high body dissatisfaction post-embodiment (insufficient data to calculate effect size) |
| Porras-Garcia et al. (2021) | 2 | *N* = 35 AN (31 females, 4 males; 16 in experimental group, 19 in control group; 2 males in each) (inpatient sample): *M* _age_ = 18.63 years; *SD* = 6.78; *M* _BMI_ = 17.48, *SD* = 1.12  Country: Spain | 1. FBI paradigm: via HMD  2. Avatars: 5 different BMIs (ranging from each participant's real-size to healthy BMI target); matched participant's height and gender  3. Subjective embodiment: ownership (FBI-VAS)  4. BID: body dissatisfaction and drive for thinness (EDI-3); body dissatisfaction and distortion (BIAS-BD); fear of gaining weight (VAS); body anxiety (VAS) | Randomly assigned to experimental or control group; both received treatment as usual (CBT, nutritional rehabilitation, and group counselling); control group received one session of FBI; experimental group received six. Avatar's BMI increased from real-size to healthy target across 5 weekly sessions. In each session, two FBI procedures: visuo-motor stimulation (movement) and visuo-tactile stimulation (touched with VR controller on legs, arms, and stomach): sync condition only. All VASs administered whilst in VR. 3-month follow-up: All BID measures. | **Subjective embodiment**  Ownership:  - Experimental group post-intervention > pre-intervention (no changes in control group; *d* = 0.43)  **BID**  Body distortion, body dissatisfaction, and fear of gaining weight:  - Reduced post-intervention and especially at follow-up for both groups; though experimental group < control group (Time*group: body distortion: *d* = 1.01; body dissatisfaction: *d* = 0.77; fear of gaining weight: *d* = 0.50)  BMI, body anxiety, and drive for thinness:  - Increased BMI (i.e., weight restoration) and reduced body anxiety and drive for thinness post-intervention and at follow-up (BMI: *d* = 0.82; body anxiety: *d* = 0.64; drive for thinness: *d* = 0.43) |
| Preston & Ehrsson (2014) | 2 | *N* = 38 (19 female, 19 male) (unspecified sample): *M* _age_ = 25.0 years; *M* _BMI_ = 21.7  Country: Sweden | 1. FBI paradigm: via HMD  2. Avatars: (1) larger body (LB) and (2) smaller/slimmer body (SB) based on participant’s hip measurements (115% and 85% adjustments, respectively)  3. Subjective embodiment: ownership (modified EQ)  4. Objective embodiment: skin conductance response  5. Objective embodiment and BID: body size estimation task (own hip width)  6. BID: body dissatisfaction (BISS: FRS); ED pathology (EDE-Q) | EDE-Q at baseline. Experiment 1: Four FBI conditions; (1) sync LB, (2) async LB, (3) sync SB, (4) async SB (order randomised across participants; 4 x trials per condition). Visuo-tactile stimulation (stroking on torso). Body size estimates at baseline. Skin conductance response measured in first three trials per condition. EQ and body size estimations in final trial per condition.  Experiment 2: Identical; however, without SCR or async conditions. BISS and FRS pre- and post-illusion. | **Subjective embodiment**  Ownership:  - Sync > async (*d’*s *=* 1.03-1.36)  **Objective embodiment**  Skin conductance response:  - Sync > async (*d* = 0.84)  **Objective embodiment and BID**  Body size estimation and body dissatisfaction:  - Pre-illusion > SB condition (body size: *d* = 2.10; body dissatisfaction: *d* = 0.70). Pre-illusion = LB condition (body size: *d* = 0.24 [pre-illusion > LB condition]; body dissatisfaction: *d* = 0.36 [LB condition > pre-illusion])  - Failed to report effects of synchrony  - Positive correlation between ED pathology and change in body satisfaction (LB only); higher scores associated with more positive changes; lower scores associated with more negative changes (LB: *d*’s = 0.89; SB: *d*’s = 0.56]. No correlation between ED pathology and change in body size estimation for SB or LB (*d*’s = 0.56-0.62; higher ED pathology associated with more negative change) |
| Preston & Ehrsson (2018) | 2 | Experiment 1:  *N* = 40 (20 males, 20 females) (unspecified sample): *M*_age_ = 27.0 years; *SD* = 5.5: *M* _BMI_ = 22.7; *SD* = 2.8  Experiment 2:  *N* = 64 (32 males, 32 females) (unspecified sample): *M*_age_ = 26.0 years; *SD* = 4.4: *M* _BMI_ = 22.2; *SD* = 2.1  Country:  Sweden | 1. FBI paradigm: via HMD  2. Avatars: sex-matched slim and obese bodies conformed with traditional ideal body types; slim male (muscular physique and BMI of 20.4), slim female (BMI of 18.4), obese male (not overtly muscular and BMI of 36) and obese female (BMI or 32.3)  3. Subjective embodiment: ownership (modified EQ)  4. BID: explicit body satisfaction (BISS); implicit body satisfaction (IAT); ED symptoms (EDE-Q) | Experiment 1: Four conditions; (1) sync; slim, (2) async; slim, (3) sync; obese, and (4) async; obese; (order randomised).  Visuo-tactile stimulation (touches on torso). BISS at baseline. BISS and EQ after each condition.  Experiment 2: identical; however, assessed IAT not BISS. | **Subjective embodiment**  Ownership:  - Sync > async (both avatar sizes; *d*’s = 1.15-1.25)  **BID**  Explicit body satisfaction:  - Slim condition > obese condition (sync > async; synchrony*avatar size: *d*’s 0.14-0.65 [obese]; *d*’s 0.05-1.04 [slim])  - Higher ED symptoms predicted greater reductions (post obese) and increases (post slim; avatar size*synchrony*ED symptoms: *d* = 0.59)  - Obese condition < baseline (sync = async; *d*’s = 0.65-0.78); Slim condition < baseline (async only; *d* = 0.55)  Implicit body satisfaction:  - Sync > async (*d* = 0.65)  - Slim condition = obese condition (*d*’s = 0.14-0.26; direction not discernible)  - Lower ED symptoms predicted greater increases post-sync relative to async (*d* = 0.73) |
| Provenzano et al. (2019) | 1, 2 | *N* = 40 females  AN group (patient sample):  *n* = 20; *M*_age_ = 23.30 years; *SD* = 7.60: *M* _BMI_ = 15.87; *SD* = 1.12  HC group (unspecified sample):  *n* = 20; *M*_age_ = 23.85 years; *SD* = 3.23: *M* _BMI_ = 18.94; *SD* = 0.98  Country: Italy | 1. FBI paradigm: via HMD  2. Avatars (-15%, 0%, +15%): (1) same body size as participant (2), 15% thinner than participant, (3) 15% fatter than participant  3. Subjective embodiment: ownership, agency, referred touch (modified EQ)  4. Objective embodiment: body temperature  5. BID: body dissatisfaction (perceived and ideal body task) | FBI: sync and async conditions for each avatar (6 x trials; order of size conditions and stimulation randomised across participants). Visuo-tactile stimulation (stroking on abdomen). Perceived/ideal body task, temperature, and EQ after each trial. | **Subjective embodiment**  Ownership, agency, and referred touch:  - Sync > async (*d*’s > 1.11)  - Ownership: Post-embodiment +15% avatar > 0% and -15% avatars (*d* = 0.83)  - AN = HC (*d* = < 0.49; direction not discernible)  **Objective embodiment**  Body temperature:  - Sync < Async (*d* = 0.28)  - AN = HC (*d* = < 0.16; direction not discernible)  **BID**  Body dissatisfaction:  - No change post-embodiment of different-sized avatars for either BID group (*d*’s < 0.49; direction not discernible) |
| Scarpina et al. (2019) | 1, 2 | *N* = 30 females  Obese group (inpatient sample):  *n* = 15; *M*_age_ = 32 years; *SD* = 6: *M* _BMI_ = 45; *SD* = 6.69  Healthy-weight control group (university and community sample):  *n* = 15; *M*_age_ = 29 years; *SD* = 8: *M* _BMI_ = 22; *SD* = 1.66  Country: Italy | 1. FBI paradigm: via HMD  2. Avatar: 25-year-old woman, thin abdomen (waist circumference was 74 cm; i.e., perceptively skinner than the obese [*M* = 140 cm] and healthy-weight [*M* = 100 cm] participants)  3. Subjective embodiment: ownership, agency, location (modified EQ)  4. Objective embodiment and BID: body size estimation task (width and circumference of own shoulders, abdomen, and hips, and overall height) | FBI: sync and async conditions (randomised order across participants; 1 x trial per condition). Visuo-tactile stimulation (stroking on abdomen). Body size estimation and EQ after each condition. Body size estimation pre-and post-assessment. | **Subjective embodiment**  Ownership, agency, and location:  - Obese group = HC group (*d* = 0.00)  - Sync > async (location only; condition*subscale: *d* = 0.62]  **Objective embodiment and BID**  Body size misestimation:  - Hip circumference: Post-embodiment skinny avatar after sync (not async) condition < baseline (obese group = HC group) (avatar size: *d* = 0.60; BID group: *d* = 0.56; obese > HC)  - Shoulder width: Baseline > post-sync (HC only; timing*BID group; *d* = 0.63)  - No effect post-embodiment for shoulder circumference, abdomen, hip width, or height (both BID groups; timing*BID group: *d*’s = 0.00-0.52) |
| Serino et al. (2016) | 2 | *N* = 21 females (university sample): *M*_age_ = 22.76 years; *SD* = 2.42: *M* _BMI_ = 21.36; *SD* = 1.91  Country: Italy | 1. FBI paradigm: via HMD  2. Avatar: skinny belly (waist circumference [74 cm] less than mean of sample [86 cm])  3. Subjective embodiment: ownership, agency, and self-location (modified EQ)  4. Objective embodiment and BID: body size estimation (width and circumference of own shoulders, abdomen, and hips, and overall height) | FBI: sync and async conditions (randomised order across participants; 1 x trial per condition). Visuo-tactile stimulation (stroking on abdomen). Body size estimation and EQ after each condition. | **Subjective embodiment**  Ownership, agency, and self-location:  - Sync > async (self-location only; no effect of synchrony for ownership and agency; synchrony*sub-scale: *d* = 0.50)  **Objective embodiment and BID**  Body size misestimation:  - Post-sync embodiment (height, abdomen width, shoulder and hip circumference) < post-async (*d*’s > 1.05). No change for shoulder/hip width or abdomen circumference (*d*’s = 0.00-0.46; post-sync > async) |
| Serino et al. (2017) | 2 | *N* = 23 females with AN (unspecified sample): *M*_age_ = 22.76 years; *SD* = 4.64: *M* _BMI_ = 15.50; *SD* = 2.14  Country: Italy | 1. FBI paradigm: via HMD  2. Avatar: skinny body (waist circumference [74 cm] less than mean of sample [86 cm])  3. Objective embodiment and BID: body size estimation (width and circumference of own shoulders, abdomen, and hips) | FBI: sync and async conditions (randomised order across participants; 1 x trial per condition). Visuo-tactile stimulation (stroking on body; no description of which parts). Body size estimation tasks at baseline and after each condition. | **Objective embodiment and BID**  Body size misestimation:  - Abdomen and hip circumference: Post-embodiment < pre-embodiment (*d*’s = 1.13-1.63)  - No effect for shoulders or abdomen and hip width (*d*’s = 0.43-1.58; post-embodiment < pre-embodiment) - Sync = async (all body parts) (insufficient data to calculate effect size) |
| Serino et al. (2020) | 2 | *N* = 26 females (university sample): *M*_age_ = 24.19 years; *SD* = 3.19: *M* _BMI_ = 20.22; *SD* = 1.27  Country: Italy | 1. FBI paradigm: via HMD  2. Avatars: (1) normal-sized (control condition), 170 cm tall; (2) enlarged, 850 cm tall; (3) shrunken, 34 cm tall  3. Subjective embodiment: ownership, agency, self-location (modified EQ)  4. Objective embodiment and BID: body size estimation (height and width of own shoulders, abdomen and hips**)** | FBI: sync condition only (3 x trials (i.e., for each avatar body size). Visuo-motor stimulation (moved hands/arms). Body size estimation and EQ after each condition. | **Subjective embodiment**  Self-location:  - Post-embodiment normal-size avatar > extreme sizes (enlarged = shrunken; *d* = 0.94)  Ownership and agency:  - No effect of body-size condition (insufficient data to calculate effect sizes)    **Objective embodiment and BID**  Body size misestimation:  - Shoulder width: underestimation post-embodiment of extremely enlarged and shrunken avatars > normal-sized (enlarged: *d* = 0.35; shrunken: *d* = 0.59)  - Abdomen and hip width: underestimation post-embodiment of shrunken avatar only > normal-sized (*d*’s = 0.29-0.41)  - Height: No effect of body-size condition (*d* = 0.49; shrunken > normal-sized > enlarged) |
| Tagini et al. (2020) | 1 | *N* = 41 (university and community sample)  Obese group:  *n* = 21; *M*_age_ = 38.67 years; *SD* = 12.45: *M* _BMI_ = 45.33; *SD* = 6.46  Healthy-weight (HC) group:  *n* = 20; *M*_age_ = 36.75 years; *SD* = 7.69: *M* _BMI_ = 22.10; *SD* = 2.82  No gender information.  Country: Italy | 1. Virtual hand illusion (variant of the classic RHI using a picture of participant’s real-size hand [displayed on screen] instead of a rubber hand)  2. Subjective embodiment: visual capture of hand position and touch (modified illusion questionnaire)  3. Objective embodiment: proprioceptive drift | Virtual hand illusion: sync and async conditions (order randomised across participants; 1 x trial per condition). Visuo-tactile stimulation (fingers stroked). Proprioceptive drift measured pre-and-post both conditions. Illusion questionnaire after each condition. | **Subjective embodiment**  Visual capture and touch:  - Sync > async (insufficient data to calculate effect size)  - HC group = obese group (sync: *d* = 0.56; async: *d* = 0.45)  **Objective embodiment**  Proprioceptive drift:  - Sync > async (*d* = 0.79)  - HC group > obese group (*d* = 1.02) |
| Tambone et al. (2021) | 2 | *N* = 30 females (university and community sample): *M*_age_ = 21.83 years; *SD* = 0.36  No overall BMI information.  Country: Italy | 1. FBI paradigm: via HMD  2. Avatars: female; (1) slim body (-30% of participant's actual body size), (2) large body (+30% of participant's actual body size) (measured via hipbones)  3. Subjective embodiment: intensity of touch, identification with avatar, and out-of-body experience (modified BOQ)  4. Objective embodiment and BID: body size estimation (hip width)  5. BID: implicit attitudes towards food based on perceived caloric content (adapted from IAT; FP-AAT); implicit attitudes towards own body, i.e., body satisfaction (BIAT) | Session 1: hip circumference measured, and BIAT and IAT/FP-AAT completed.  Sessions 2 and 3: FBI with slim or large avatar (order randomised across participants; 1 x trial per condition). Visuo-tactile stimulation (vibration on abdomen). BOQ and all measures from session 1 at end of each session. | **Subjective embodiment**  Touch, identification, and out-of-body experience:  - Embodiment occurred irrespective of avatar's body size (insufficient data to calculate effect sizes)  **Objective embodiment and BID**  Body size misestimation:  - Underestimation post-slim condition (*d* = 0.57); overestimation post-large condition (*d* = 0.47)  **BID**  Implicit attitudes:  - Avoidance of high-calorie food: Post-slim condition > baseline (*d* = 0.39); no effect post-large condition (insufficient data to calculate effect size)  - Body satisfaction: Not modulated by avatar size (insufficient data to calculate effect size) |
| Themelis et al. (2021) | 2 | *N* = 24 males (university and community sample): *M*_age_ = 24.00 years; *SD* = 4.95: *M* _BMI_ = 24.00; *SD* = 4.00  Country: Australia | 1. Body re-sizing illusion paradigm (no clear description; presumably followed same principles as the FBI, however, whilst wearing a HMD, participants view real-time video of their own back)  2. Subjective embodiment: ownership, agency, and feelings towards own back (modified EQ)  3. Objective embodiment and BID: back-size/shape estimation task (own shoulder and hip width)  4. BID: perceived self-capacity of the back, i.e., back strength, lifting confidence, and back fitness (VAS) | Illusion: sync condition only (4 x trials; order randomised; (1) Normal size [no visual manipulation; control condition]; (2) Strong [overlaid image of generic muscled back, shoulders 25% wider, hips 25% narrower]; (3) Reshaped [same as strong condition without overlaid image]; (4) Large [shoulders and hips 25% wider]. Visuo-motor synchrony (lifting tasks). Back size estimation at baseline. EQ, VAS, and back size estimation after each condition. | **Subjective embodiment**  Ownership and agency:  - Normal > Strong; Reshaped > Strong; Large > Strong (*d’s* = 2.21-3.26)  Feelings towards own back:  - No effect of size condition (*d* = 0.30; Reshaped > Normal > Strong and Large)  **Objective embodiment and BID**  Back size/shape misestimation:  - Shoulder and shoulder/hip (consistent with the direction of manipulation): Post-Reshaped condition > Normal (shoulder only) and Large (both measures) conditions (*d* = 0.77 [shoulder]; *d* = 1.06 [shoulder/hip])  - Hip: no effect of condition (*d* = 0.30; Large > Strong > Reshaped > Normal)  **BID**  Perceived self-capacity of the back:  - No effect of condition (*d* = 1.07; generally, Strong and Normal > Reshaped and Large) |
| Wolf et al. (2021) | 2 | *N* = 52 females (university sample) (26 per condition:  embodiment; *M* _BMI_ = 22; *SD* = 2.2: no embodiment; *M* _BMI_ = 21.8; *SD* = 3)  No age information.  Country: Germany | 1. FBI paradigm: via HMD  2. Avatar: female; BMI of 22.25  3. Subjective embodiment: ownership (VEQ)  4. Objective embodiment and BID: body-weight estimation task (estimated avatar’s BMI: research suggests that embodiment might contribute to attribution of one’s body weight to avatar’s perceived body weight) | FBI: embodiment (1PP; sync) and no-embodiment conditions (3PP; only observed avatar move) (order randomised across participants). Visuo-motor stimulation (5 x tasks involving hand, arm, hip, and leg movements).  VEQ in virtuo. Estimated avatars body weight and VEQ post-experience. | **Subjective embodiment**  Ownership:  - In virtuo: Embodiment > no embodiment (*d* = 1.46)  - Post-experience: Embodiment = no embodiment (*d* = 0.32; embodiment > no-embodiment)  Agency:  - Embodiment > no embodiment (in virtuo and post-experience; *d*’s = 2.14-2.98)  **Objective embodiment and BID**  Body-weight estimation:  - Post ‘embodiment’ condition < ‘no embodiment’ condition (*d* = 0.68)  - Embodiment (but not ‘no embodiment’) biased estimation with an increased difference between one’s own and the avatar’s BMI (BMI*condition; *d* = 0.98) |
| Zopf et al. (2016) | 1 | *N* = 46 females  AN group (inpatient sample): *n =* 23*: M*_age_ = 21.87 years; *SD* = 2.79: *M* _BMI_ = 15.82; *SD* = 1.27  HC group (university sample): *n =* 23*: M*_age_ = 21.48 years; *SD* = 2.35: *M* _BMI_ = 21.16; *SD* = 2.10  Country: Australia | 1. RHI paradigm  2. Subjective embodiment: ownership, agency, location; (modified RHI-Q)  3. Objective embodiment: reaching task  4. BID: ED psychopathology (EDI-3; BSQ; BPSS) | RHI: sync and async conditions (order randomised across participants; 2 x trials per condition). Visuo-tactile stimulation (hands stroked). Reaching task (15 randomly presented trials) and RHI-Q after each trial. | **Subjective embodiment**  Ownership, agency, and self-location:  - Sync > async (*d’*s = 1.22-1.42)  - AN > HC (*d* = 0.61)  - No correlation with ED psychopathology (insufficient data to calculate effect sizes)    **Objective embodiment**  Reaching task:  - AN > HC (*d* = 0.61)  - Sync > async (*d* = 1.34)  - No correlation with ED psychopathology (insufficient data to calculate effect sizes) |

*Note*: 1PP = first person perspective; 3PP = third person perspective; AN = anorexia nervosa; async = asynchronous; BAS = Body Appreciation Scale; BCI = Body Change Inventory; BDD = body dysmorphic disorder; BED = binge eating disorder; BIAS-BD = Body Image Assessment Scale-Body Dimensions; BIAT = Body Brief Implicit Association Test; BISS = Body Image States Scale; BMI = body mass index (kg/m^2^); BN = bulimia nervosa; BPSS = Body Parts Satisfaction Scale; BOI-Q = Body Ownership illusion questionnaire; BOQ = Body Ownership questionnaire; BSS = Body Satisfaction Scale; BSQ = Body Shape Questionnaire; DCQ = Dysmorphic Concern Questionnaire; ED = eating disorder; EDE-Q = Eating Disorder Examination Questionnaire; EDNOS = eating disorder not otherwise specified; EDI = eating disorder inventory; EQ = Embodiment Questionnaire; FBI = full body illusion; FP-AAT = Food Preferences Approach-Avoidance Test; FRS = Figure Rating Scale; HC = healthy control; HMD = head mounted display; IAT = Implicit Associations Task; ICD = International Classification of Diseases; *IQR* = interquartile range; *M* = mean; *Mdn* = median; MINI = Mini-International Neuropsychiatric Interview; OBCS = Objectified body consciousness scale; OSFED = other specified feeding and eating disorder; PASTAS = Physical Appearance State and Trait Anxiety Scale; REC = recovered from eating disorder; RHI = rubber hand illusion; RHI-Q = Rubber Hand Illusion Questionnaire; RRT = Relational Responding Task; sync = synchronous; VAS = visual analogue scale; VEQ = Virtual Embodiment Questionnaire; VR = virtual reality.

*Unless otherwise stated, findings reported are based upon quantitative statistics (i.e., *p* < .05 or adjusted if involved multiple comparisons) and involve main effects.

The direction of effects for non-significant findings are reported after the effect size statistic (note, the direction was only presented for non-negligible effects, i.e., *d* > 0.00).
